# Mechanistic insights into microbiome-dependent and personalized responses to dietary fibre in a randomized controlled trial

**DOI:** 10.1101/2025.11.20.25340625

**Authors:** Anissa M. Armet, Fuyong Li, Edward C. Deehan, Daria D. Nikolaeva, Omar Delannoy-Bruno, Léa Siegwald, Bernard Berger, Kaori Minehira Castelli, Dmitry A. Rodionov, Aleksandr A. Arzamasov, Junhong Liu, Benjamin Seethaler, Janis L. Cole, Khoi Nguyen Nguyen, Mingliang Jin, Yuan-Yuan Zhao, Arya M. Sharma, Jonathan M. Curtis, Spencer D. Proctor, Stephan C. Bischoff, Wendy V. Wismer, Andrei L. Osterman, Jeffrey A. Bakal, Russell Greiner, Catherine J. Field, Dan Knights, Carla M. Prado, Jens Walter

## Abstract

Dietary fiber supplementation can reduce cardiometabolic risk, but its effective use is limited by incomplete understanding of fibre-microbiome interactions and highly individualized responses. We tested acacia gum (AG; fermentable fibre), resistant starch type 4 (RS4; fermentable fibre), and microcrystalline cellulose (MCC; non-fermentable control fibre) in a six-week randomized trial in adults with excess body weight. Multi-omics profiling revealed distinct, structure-specific microbiota and short-chain fatty acid shifts with AG and RS4, which were not directly linked to physiological outcomes. Improvements in inflammation, gut barrier function, and satiety occurred across all arms, indicating fermentation-independent effects. AG reduced plasma ghrelin, linked to microbial carbohydrate-active enzyme genes targeting its structures. Machine-learning models predicted individualized, fiber-specific effects on blood pressure (AG) and C-reactive protein (RS4) from microbial pathways and fecal bile acids. These findings delineate fermentation-dependent and independent mechanisms of fibre action and provide a mechanistic basis for personalized fibre supplementation.

**Trial registration:** ClinicalTrials.gov NCT02322112

## INTRODUCTION

Dietary fibre comprises carbohydrates – including non-starch polysaccharides, oligosaccharides, and resistant starches – that are not digested by enzymes in the human small intestine. Epidemiological studies provide strong evidence that high intakes (>25 g/d) of intrinsic fibre found in whole-plant foods reduce risk of several chronic diseases, such as type 2 diabetes ^1–3^, cardiovascular disease ^1,4,5^, and overall mortality ^1^. The cardiometabolic benefits of fibre are elicited through a variety of mechanisms. For example, intrinsic fibres form the three-dimensional cell-wall matrix of whole-plant foods, delaying nutrient absorption due to reduced accessibility ^6^. Viscous fibres form gel-like substances in the gut that reduce nutrient absorption and blunt postprandial glycemia and lipidemia ^7,8^. In addition, dietary fibres influence bile acid profiles, either through direct binding or indirectly through microbiome-mediated modulation ^9^.

Many fibres are, at least to some degree, fermentable by colonic microbiota, providing growth substrates to microbes that possess the enzymatic capabilities to utilize specific structures ^9^. Such fibres modulate microbiome composition over both shorter ^10,11^ and longer ^12,13^ timeframes, and they are often referred to as prebiotics, enriching for putatively beneficial microbial taxa in a highly structure-specific manner ^14^. Fibre fermentation produces bioactive microbial metabolites, such as short-chain fatty acids (SCFAs), which modulate immunometabolic pathways implicated in cardiometabolic diseases ^15^. For example, SCFAs stimulate appetite hormones (*e.g.,* glucagon-like peptide-1 [GLP-1], peptide tyrosine tyrosine [PYY]) that promote satiety and improve insulin sensitivity ^16^. Further, SCFAs can increase the expression of tight-junction proteins and epithelial mucus production to enhance gut barrier function ^17^, and modulate immune cell function (*e.g.,* induction of Treg cells) to attenuate pro-inflammatory cytokine production ^18^, both of which can reduce inflammation ^15^. Fermentable fibres are also known to influence microbial bile acid transformation, leading to the formation of signalling molecules that influence processes such as insulin metabolism ^19^.

Mechanisms by which fibre intake can benefit health are well-established in animal models, wherein the gut microbiome makes clear causal contributions to physiological effects ^20,21^. In clinical intervention studies, benefits of fibre have been correlated with, and even predicted by, baseline microbiome features or fibre-induced shifts in the microbiome ^22,23^, with SCFAs potentially mediating these effects ^24^. However, it has been difficult to make direct causal inferences in human trials ^14^; it is, therefore, still unclear to what degree microbial fermentation contributes to clinical benefits of fibre and what the mechanisms are by which fibre-microbiome interactions influence pathophysiological processes. This is directly relevant to understand the clinical effects of isolated and synthetic fibre supplements, which are much more variable and studies often inconclusive when compared to intrinsic fibre ^25–27^. There are many potential explanations for this discrepancy (*e.g.,* lack of three-dimensional cell-wall matrix and associated phytochemicals, lower structural complexity, lower dose) ^28^, and effects of supplements are, therefore, likely more specific and dependent on the physicochemical properties of the particular fibre used ^29^. However, if fermentation would be relevant, the health effects of a specific fibre chemistry might also depend on whether an individual’s microbiome possesses the genetic capacity to degrade the structure. Microbiome heterogeneity may, therefore, contribute to the personalized clinical responses observed with dietary fibre supplements ^30–32^.

Fibre supplementation is attracting growing interest for its translational potential, both in reformulating processed foods and in developing microbiome-targeted ^33,34^ and personalized ^35,36^ dietary strategies. Yet translation is hindered by key knowledge gaps: the extent to which microbial fermentation mediates the clinical benefits of fibre, the mechanisms by which distinct fibre structures interact with the microbiome to influence host physiology, and whether inter-individual variation in microbiome composition and functional capacity explains variable clinical responses. To address these questions, we conducted a randomized controlled trial in adults with excess body weight, testing the immunometabolic effects of three structurally-distinct fibres with different fermentability profiles. We applied multi-omics profiling to characterize taxonomic and functional features of the gut microbiome alongside mechanistic markers of host-microbiome interactions and cardiometabolic risk markers. Finally, we applied machine learning to assess whether fibre-induced microbiome shifts associate with physiological effects of the fibres, and if clinical responses can be predicted through baseline microbiome features (primary outcome).

## RESULTS

### A randomized controlled trial to characterize physiological effects of structurally-distinct fibres in adults with excess body weight

We performed a six-week, parallel-arm, randomized controlled trial in adults with excess body weight (body mass index [BMI] 25-35kg/m^2^) and compared two structurally-distinct fermentable fibres – acacia gum (AG) and resistant starch type IV (RS4), to an unfermentable microcrystalline cellulose (MCC) control fibre (**Fig. 1a**). AG primarily consists of type II arabinogalactan chains, containing a β-(1,3) galactose backbone decorated with β-(1,6) galactose linked to α-(1,3) and T-arabinose residues, in addition to T-rhamnose and, to a lesser extent, β-(1,6) and T-glucuronic acid residues ^37–39^. AG is water-soluble, fermentable, and has low-viscosity ^40^. The RS4 used in this trial was a phosphorylated, cross-linked, wheat-based resistant starch. RS4 consists of a linear α-(1,4)-linked glucose backbone with α-(1,6)-linked branch points, is water-insoluble, fermentable, and has low-viscosity ^41^. MCC consists of a linear β-(1,4) glucose backbone, and is non-fermentable, water-insoluble, and non-viscous ^42^. Participants were provided individual daily sachets (25 g/d for females, 35 g/d for males) of their assigned fibre to incorporate into their usual diet for six weeks, with a two-day run-in period wherein half-doses (12.5 or 17.5 g/d for females and males, respectively) were provided.

**Fig. 1.**
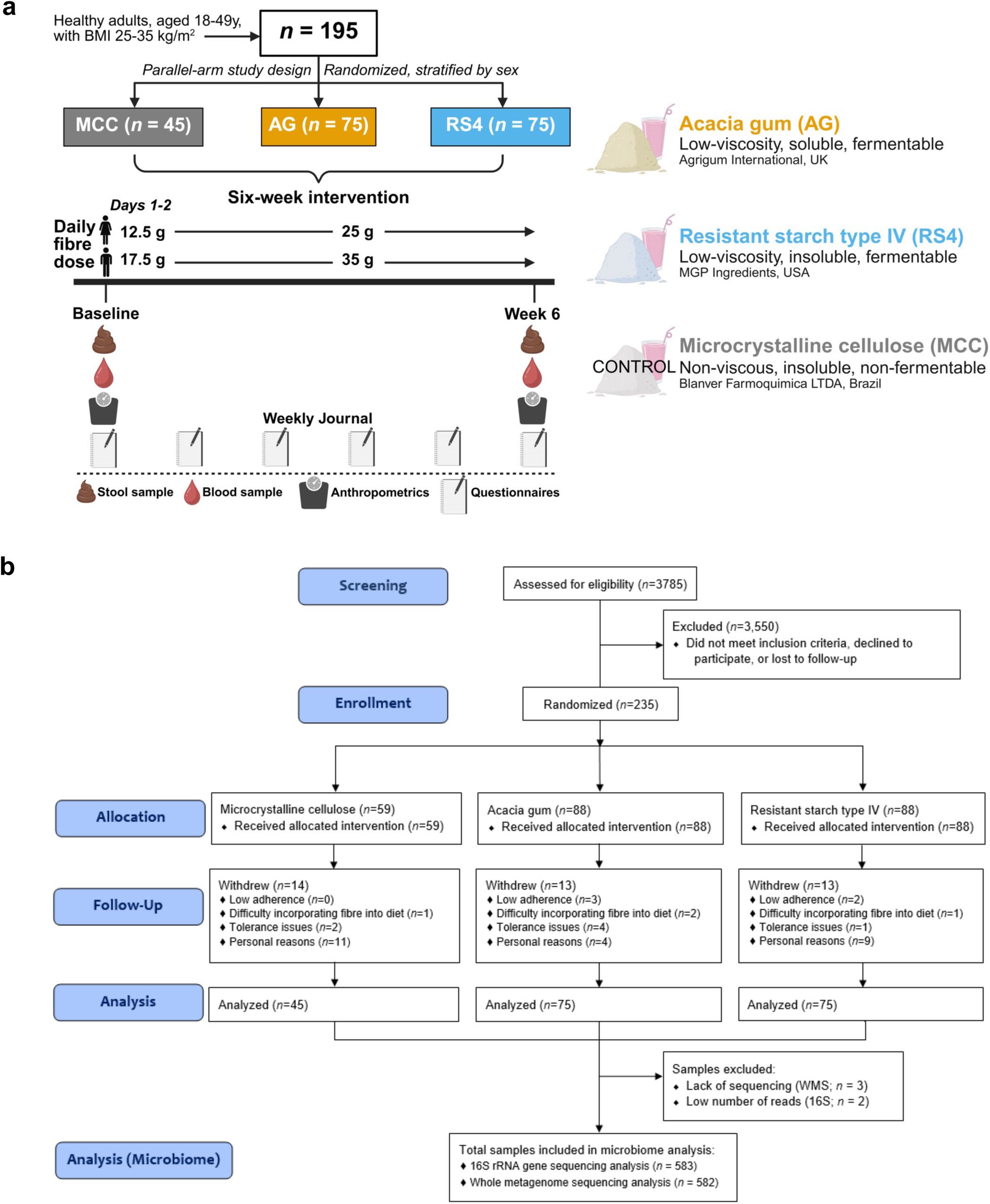
Study design and participant flow diagrams. **a**, Depiction of randomized controlled trial design and sample type collection. **b**, CONSORT diagram of the study flow. 16S, 16S rRNA gene amplicon sequencing; AG, acacia gum; BMI, body mass index; MCC, microcrystalline cellulose; RS4, resistant starch type IV; WMS, whole metagenome sequencing.

Between August 2015 and April 2020, over 3,700 individuals expressed interest in participating and were screened for eligibility, 235 were randomized to one of the three arms, and 195 completed the six-week intervention (**Fig. 1b**). Average age and BMI were not different between study groups, with baseline characteristics reported in **Table 1**. Adherence to the intervention was over 95% in each arm, and no serious adverse events related to the study were reported.

**Table 1.** Baseline characteristics.

|  | All participants<br>(n = 195) | AG<br>(n = 75) | RS4<br>(n = 75) | MCC<br>(n = 45) |
| --- | --- | --- | --- | --- |
| **Demographics** |  |  |  |  |
| Sex (female/male) | 132/63 | 51/24 | 51/24 | 30/15 |
| Age (years) | 31 (12) | 31 (12) | 31 (10) | 30 (13) |
| **Risk markers of chronic diseases** |  |  |  |  |
| Body weight (kg) | 81.9 (19.4) | 84.2 (21.2) | 81.7 (19.6) | 81.1 (18.3) |
| Body mass index (kg/m<sup>2</sup>) | 28.5 (4.0) | 28.6 (4.2) | 28.5 (4.1) | 28.0 (4.2) |
| Waist circumference (cm) | 93.1 (11.4) | 93.3 (10.1) | 92.9 (13.9) | 92.4 (10.4) |
| Body fat (%) | 33.7 (11.7) | 35.0 (11.4) | 33.3 (11.0) | 33.6 (11.5) |
| Total cholesterol (mmol/L) | 4.7 (1.1) | 4.8 (1.2) | 4.7 (1.1) | 4.7 (1.1) |
| LDL (mmol/L) | 2.8 (1.0) | 3.0 (1.1) | 2.7 (0.9) | 2.6 (0.7) |
| HDL (mmol/L) | 1.3 (0.5) | 1.3 (0.4) | 1.4 (0.5) | 1.3 (0.5) |
| Non-HDL (mmol/L) | 3.3 (1.1) | 3.5 (1.4) | 3.3 (1.0) | 3.2 (0.9) |
| Triglycerides (mmol/L) | 1.1 (1.0) | 1.2 (1.0) | 1.0 (0.9) | 1.0 (0.9) |
| Systolic blood pressure (mmHg) | 119.5 (16.8) | 120.5 (18.5) | 119.5 (15.0) | 119.0 (18.5) |
| Diastolic blood pressure (mmHg) | 74.5 (12.0) | 74.5 (11.3) | 75.5 (12.8) | 75.0 (10.0) |
| Heart rate | 73.0 (12.3) | 73.5 (11.0) | 71.0 (12.3) | 75.0 (10.0) |
| Glucose (mmol/L) | 4.9 (0.5) | 4.9 (0.5) | 5.0 (0.5) | 4.9 (0.5) |
| QUICKI | 0.37 (0.04) | 0.37 (0.04) | 0.38 (0.04) | 0.37 (0.04) |
| Insulin (μIU/L) | 5.5 (3.3) | 5.8 (3.5) | 5.1 (2.9) | 5.4 (3.4) |
| HOMA-IR | 1.2 (0.7) | 1.3 (0.7) | 1.1 (0.7) | 1.2 (0.7) |
| C-reactive protein (mg/L) | 1.9 (2.8) | 2.5 (2.7) | 1.9 (2.2) | 1.5 (2.8) |
| **Mechanistic markers of host-microbiome interactions** |  |  |  |  |
| Fecal calprotectin (ng/mg) | 37.5 (45.9) | 39.5 (35.3) | 32.5 (44.8) | 50.2 (71.0) |
| Plasma LBP (μg/mL) | 6.0 (4.1) | 5.9 (3.8) | 5.7 (4.4) | 6.6 (3.9) |
| Fecal zonulin (ng/mg) | 107.8 (134.5) | 103.3 (126.3) | 107.8 (137.4) | 114.0 (152.5) |
| Ghrelin (pg/mL) | 278.4 (207.8) | 267.7 (182.6) | 287.0 (216.8) | 276.7 (187.8) |
| Peptide YY (pg/mL) | 29.9 (15.8) | 29.1 (15.8) | 30.3 (16.0) | 29.9 (14.4) |
| Leptin (pg/mL) | 4312.5 (6028.9) | 4323.8 (6315.5) | 3757.6 (4778.6) | 4938.2 (8309.7) |
| Glucagon-like peptide 1 (pg/mL) | 0.66 (0.57) | 0.68 (0.61) | 0.59 (0.56) | 0.62 (0.41) |
| Glucagon (pg/mL) | 19.4 (10.8) | 19.2 (12.8) | 18.5 (9.0) | 21.2 (11.6) |
| Adiponectin (pg/mL) | 12.8 (7.8) | 11.9 (5.6) | 14.2 (7.6) | 12.3 (9.2) |
| Tumor necrosis factor-α (pg/mL) | 0.50 (0.15) | 0.51 (0.16) | 0.49 (0.15) | 0.54 (0.11) |
| Interleukin-6 (pg/mL) | 0.31 (0.20) | 0.31 (0.22) | 0.26 (0.21) | 0.34 (0.15) |
| Interleukin-8 (pg/mL) | 1.5 (0.6) | 1.5 (0.6) | 1.5 (0.8) | 1.5 (0.6) |
| Interleukin-10 (pg/mL) | 0.14 (0.09) | 0.14 (0.10) | 0.16 (0.09) | 0.14 (0.06) |
| Trimethylamine-N-oxide (μM) | 2.2 (1.8) | 2.4 (1.7) | 2.3 (2.1) | 1.8 (1.4) |

Data are presented as median (interquartile range).
AG, acacia gum; HDL, high-density lipoprotein cholesterol; HOMA-IR, homeostatic model assessment of insulin resistance; LBP, lipopolysaccharide binding protein; LDL, low-density lipoprotein cholesterol; MCC, microcrystalline cellulose; QUICKI, quantitative insulin sensitivity check index; RS4, resistant starch type IV.

### Effects of fibre supplements on markers of transit time and gastrointestinal symptoms

The fibre supplements did not alter markers of transit time – self-reported stool consistency (**Fig. S1a**) or bowel movement frequency (**Fig. S1b**). Non-fermentable MCC did not induce gastrointestinal symptoms throughout the intervention (**Fig. S1c-f**). RS4 led to a small but statistically significant increase in stomach-aches (**Fig. S1c**) and bloating (**Fig. S1d**), and both AG and RS4 increased flatulence (**Fig. S1e**) and overall symptom scores (**Fig. S1f**) compared to baseline (all FDR-adjusted *p*<0.05, generalized estimating equations with repeated-measures models). However, reported symptoms were apparently reduced towards the end of the intervention, both visually and based on reduced levels of significance (**Fig. S1c-f**). This may indicate adaptation towards improved tolerance to the fermentable fibres over six weeks, consistent with findings in our previous study on arabinoxylan ^43^.

**Fig. S1.**
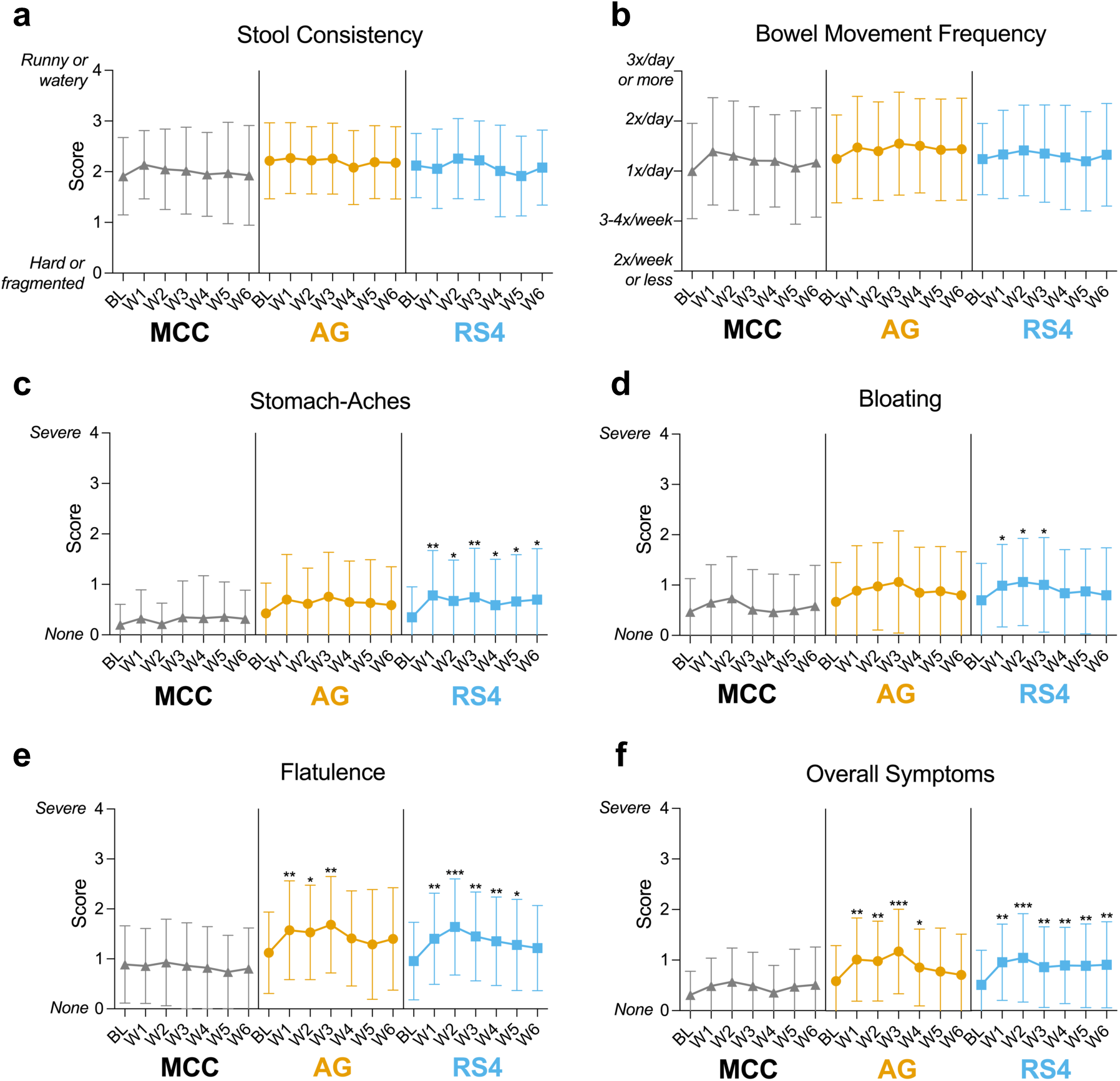
Effects of fibre supplementation on markers of transit time and gastrointestinal tolerance. Self-reported **a**, stool consistency, **b**, bowel movement frequency, and **c-f**, gastrointestinal symptoms were assessed via questionnaire weekly during the six-week intervention. Data are presented as mean ± SD. * = *p* < 0.05; ** = *p* < 0.01; *** = *p* < 0.001; generalized estimating equations with repeated-measures models. AG, acacia gum; BL, baseline; MCC, microcrystalline cellulose; RS4, resistant starch type IV; W, week.

### Impact of fermentable fibres on overall gut microbial profiles

The gut microbiome was characterized using both 16S rRNA gene amplicon sequencing and shotgun whole metagenome sequencing (WMS). For the WMS dataset, two complementary approaches were applied: reference-mapping-based profiling (*i.e.*, reference-based) and *de novo* assembly-based profiling (*i.e.*, metagenome-assembled-genome [MAG]-based).

Compared to baseline, MCC had no effects on α-diversity metrics of compositional and functional features (**Fig. S2a-c**), while both AG and RS4 led to modest but significant reductions in α-diversity indices compared to baseline and the MCC group based on compositional features (*e.g.,* MAGs, amplicon sequence variants [ASVs], and reference-based species), including decreases in the number of observed features (**Fig. S2a**), Pielou’s evenness (**Fig. S2b**), and Shannon index (**Fig. S2c**) (all FDR-adjusted *p*<0.05; paired Wilcoxon tests for within-group analyses, unpaired Wilcoxon tests for between-group analyses). These findings align with previous studies on fermentable fibres ^33,42^, which have reported similar decreases. Further, both AG and RS4 significantly altered overall gut compositional structures compared to MCC, as demonstrated by increased Bray-Curtis distances between baseline and the end (week 6) of the intervention (intra-individual differences, estimated based on MAGs, ASVs, and reference-based species), with AG exerting a stronger effect compared to RS4 (all FDR-adjusted *p*<0.05) (**Fig. S2d**).

**Fig. S2.**
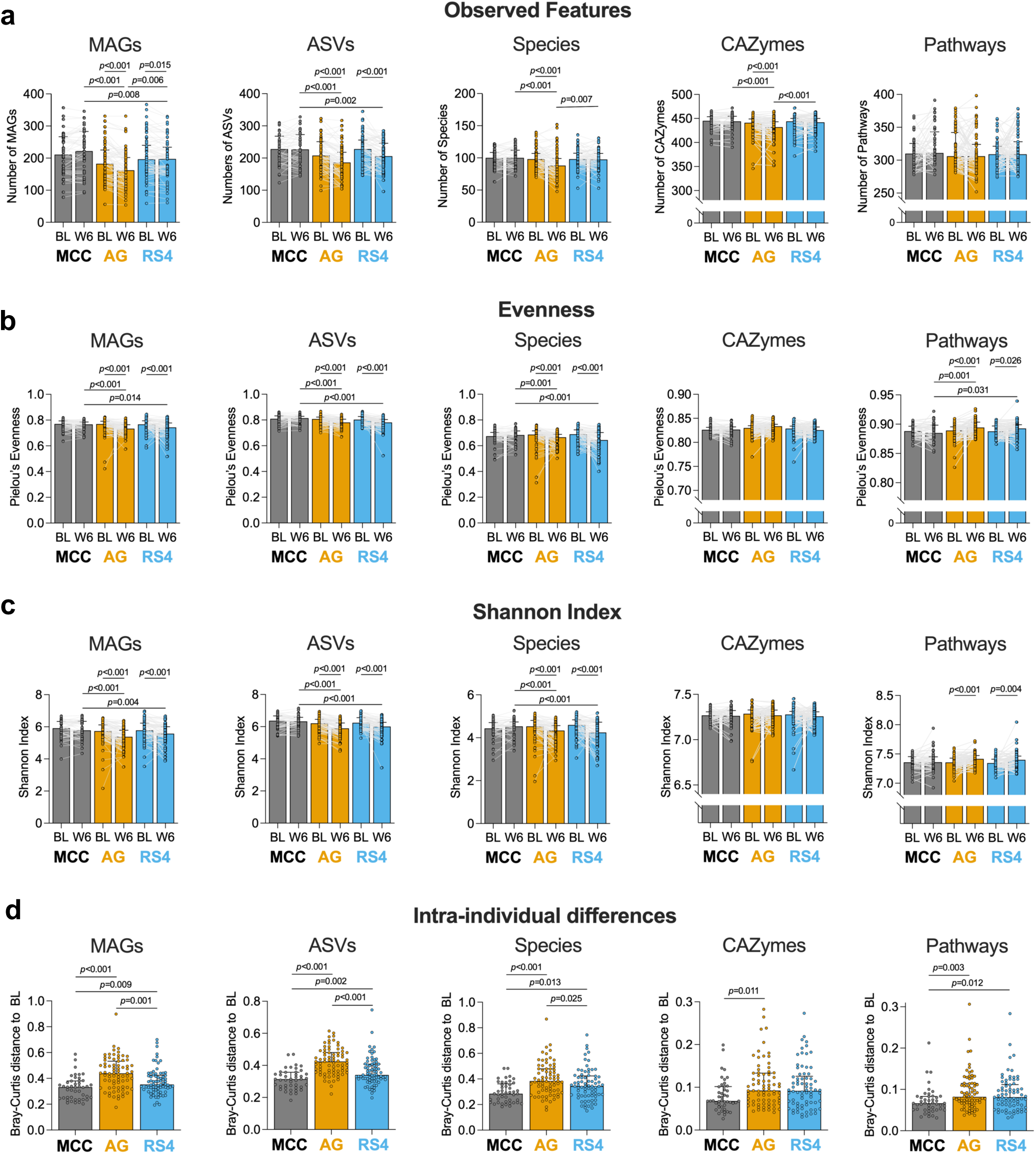
Changes to diversity indices as a result of fibre supplementation. Bar plots show intra-individual diversity indices for compositional and predicted functional features of the gut microbiome. **a,** Observed features, **b,** Pielou’s evenness, **c,** Shannon index, and **d**, Bray-Curtis dissimilarity of each participant’s microbiome between baseline and Week 6 (*i.e.,* intra-individual *β*-diversity). Symbols represent individual participant changes, data are presented as median ± IQR. FDR-adjusted *p* < 0.05 considered significant (within-group analyses – paired Wilcoxon tests; between-group analyses – unpaired Wilcoxon tests). AG, acacia gum; ASVs, amplicon sequence variants; BL, baseline; CAZymes, carbohydrate-active enzymes; MAGs, metagenome assembled genomes; MCC, microcrystalline cellulose; RS4, resistant starch type IV; W6, week 6.

In contrast to the reduced α-diversity observed in compositional profiles, both AG and RS4 increased the evenness (**Fig. S2b**) and Shannon index (**Fig. S2c**) of reference-based pathways (FDR-adjusted *p*<0.05), suggesting a potential functional diversification despite compositional contraction in response to the fermentable fibres. Additionally, AG, but not RS4, significantly reduced the number of observed CAZymes (FDR-adjusted *p*<0.001) (**Fig. S2a**), while the evenness (**Fig. S2b**) and Shannon index (**Fig. S2c**) of CAZymes remained unchanged during the intervention. Overall, these results indicate that the fermentable fibres reduce α-diversity of the gut microbiome composition, yet their impact on microbiome functions is less pronounced and differs between metabolic pathways and carbohydrate-utilization capacities.

### Fermentable fibres induce highly specific changes in the relative abundances of microbiome compositional and functional features

Consistent with its non-fermentable properties, MCC had no detectable effect on the abundance of microbial taxa (all taxa from MAG-based, reference-based, and 16S rRNA gene amplicon sequencing; FDR-adjusted *p*>0.1 for paired [within-group] Wilcoxon tests; Supplementary Table 1). In contrast, AG and RS4 induced highly specific changes in the relative abundances of microbial taxa (Supplementary Table 1), with no overlap in the species that were enriched. AG selectively increased the average relative abundances of several species, including *Bifidobacterium longum* (MAG027)*, Gemmiger formicilis* (MAG094)*, Blautia obeum* (MAG055)*, Lachnospiraceae* KLE1615 sp. (MAG088), and *Faecalibacterium prausnitzii* (MAG110) (**Fig. 2a**; FDR-adjusted *p*<0.01 for both within-group and between-group comparisons with MCC; Wilcoxon tests). In contrast, compared to MCC, RS4 increased the average relative abundances of *Bifidobacterium adolescentis* (MAG077), *Parabacteroides distasonis* (MAG206)*, Acutalibacteraceae* UBA1417 sp. (MAG111), *Eisenbergiella* sp. (MAG040), and *Anaerostipes hadrus* (MAG067). These compositional changes were overall consistent with changes identified through ASV-based (**Fig. S3a**) and reference-based (**Fig. S3b**) analyses (Supplementary Table 1). While reductions in the abundances of bacterial taxa were mostly specific to AG and RS4, both fibres led to significant decreases in the abundances of *Mediterraneibacter torques* (MAG238) (synonym of *Ruminococcus torques*), a known mucin degrader ^44^, and *Mediterraneibacter faecis* (MAG068) (FDR-adjusted *p*<0.01; **Fig. 2a**). Importantly, the effects of RS4 on microbial taxa were consistent with our previous studies across different cohorts and sequencing platforms, including enrichments of *Bifidobacterium adolescentis*, *Parabacteroides distasonis*, and *Eisenbergiella sp.*, and the reduction in *Ruminococcus torques* ^33,45^. Collectively, these findings underscore that structurally-distinct fermentable fibres can drive highly precise and selective shifts in gut microbial taxa ^33,45^.

**Fig. 2.**
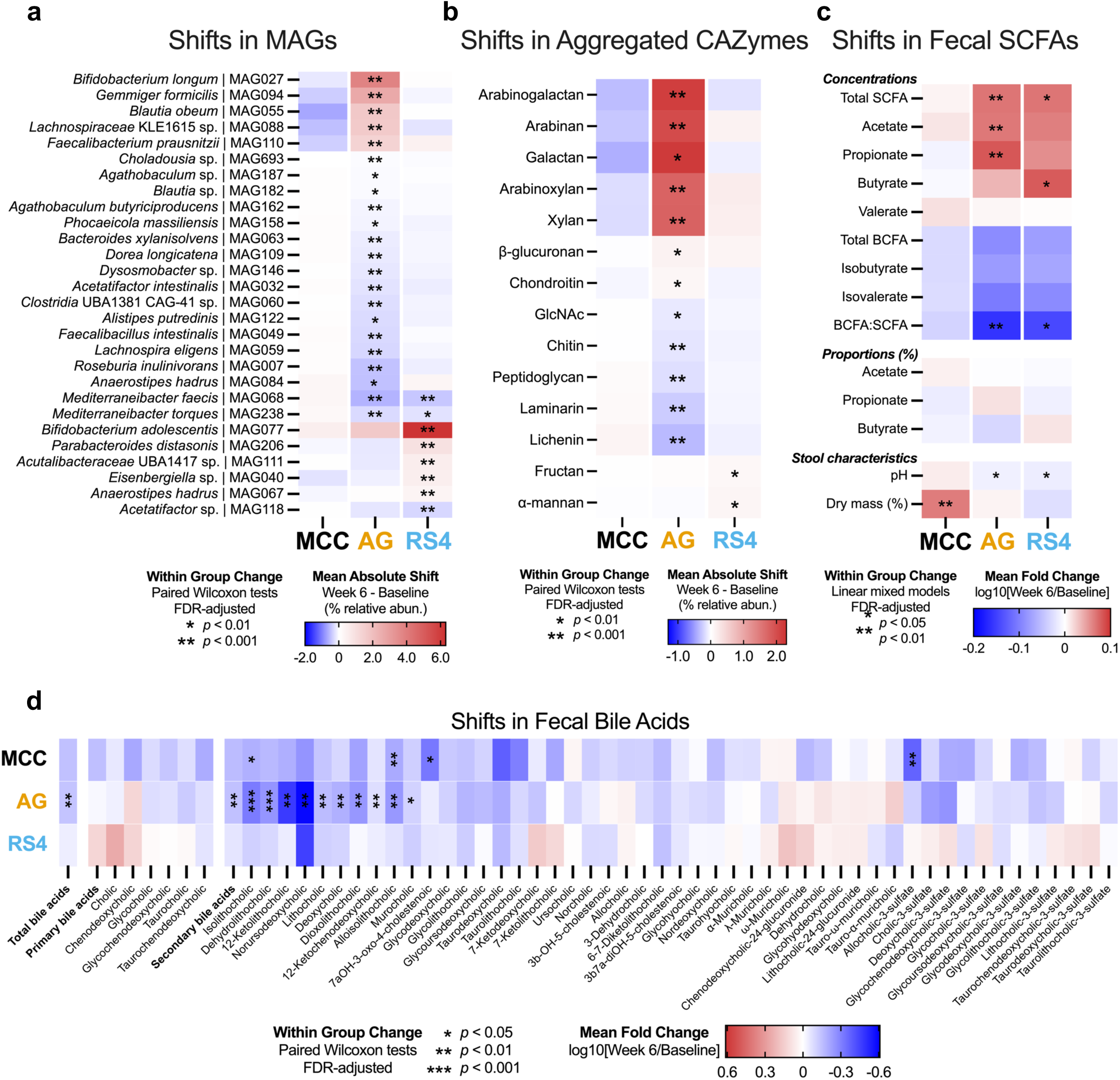
Selective effects of structurally-distinct fibres on gut microbiome compositional and functional features. Mean absolute shifts from baseline to week 6 in relative abundances of **a**, MAGs, and **b**, aggregated CAZymes. Mean log_10_ fold changes from baseline to week 6 in fecal **c**, SCFAs, and **d**, bile acids. Features identified in **a** are those that were significantly altered within each fibre group (paired Wilcoxon tests; FDR-adjusted *p* < 0.01) and significantly different compared to MCC (unpaired Wilcoxon tests on absolute shifts; FDR-adjusted *p* < 0.01). The same statistical tests were applied to the aggregated abundances of CAZymes and bile acid data (FDR-adjusted *p* < 0.05), while linear mixed models were applied to the SCFA data (FDR-adjusted *p* < 0.05). AG, acacia gum; CAZymes, carbohydrate-active enzymes; MAGs, metagenome assembled genomes; MCC, microcrystalline cellulose; RS4, resistant starch type IV; SCFAs, short-chain fatty acids.

MCC also did not significantly affect microbiome functional features, such as CAZymes and metabolic pathways (all FDR-adjusted *p*>0.1 for paired [within-group] Wilcoxon tests; Supplementary Table 1). Fermentable fibres altered the relative abundances of predicted functional features, but effects were less structure-specific compared to effects on taxonomic composition (Supplementary Table 1). The relative abundances of numerous microbial pathways (81 out of 218 identified reference-based pathways) were impacted by both AG and RS4 (*e.g.,* increased PWY-5384: sucrose degradation IV [sucrose phosphorylase] and P124-PWY: Bifidobacterium shunt) (**Fig. S3c**). While some overlap was observed in the CAZymes that were altered by both AG and RS4 (**Fig. S3d**), differences emerged that reflected the distinct glycan compositions and structures of these two fibres. Specifically, the glycoside hydrolase (GH)-43 family and its subfamilies (*e.g.,* GH43_18, GH43_19, GH43_22, GH43_23, GH43_34), which potentially target arabinogalactan, were only significantly enriched in the AG group (**Fig. S3d**, Supplementary Table 1). This specificity was also reflected in the abundance of aggregated CAZymes (CAZyme subfamilies that are known or predicted to target the same glycan) that target arabinogalactan, arabinan, and galactan all increased in the AG group (**Fig. 2b**). RS4 supplementation increased the abundance of some GH13 subfamilies (*e.g.,* GH13_3, GH13_15; Supplementary Table 1), which are involved in starch metabolism. Interestingly, the relative abundance of aggregated CAZymes targeting starch was not altered by RS4; rather, those targeting fructan and α-mannan were significantly increased (**Fig. 2b**). Overall, these results highlight that AG and RS4 led to highly specific modulations of the gut microbiota at the taxonomic level, accompanied by parallel changes in CAZymes that were, although less specific, targeted against glycan substrates present in the fermentable fibres.

**Fig. S3.**
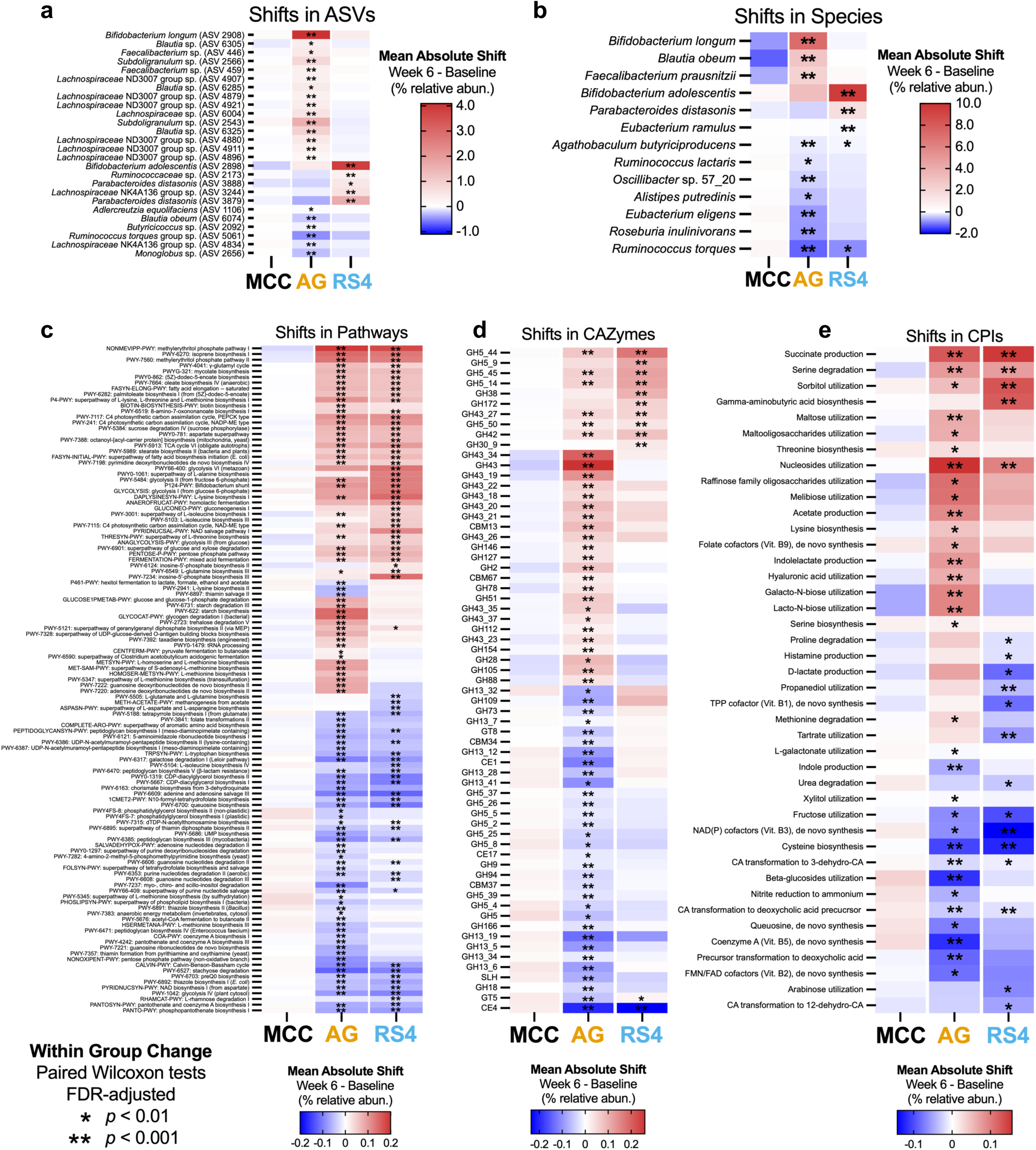
Selective effects of fermentable fibres on gut microbiome composition and functional potential. Mean absolute shifts from baseline to week 6 in relative abundances of **a**, ASVs, **b**, species, **c**, pathways, **d**, CAZymes, and **e**, CPIs. Features presented in **a-d** are those that were significantly altered within each fibre group (paired Wilcoxon tests; FDR-adjusted *p* < 0.01) and significantly different compared to MCC (unpaired Wilcoxon tests on absolute shifts; FDR-adjusted *p* < 0.01). AG, acacia gum; ASVs, amplicon sequence variants; CA, cholic acid; CAZymes, carbohydrate-active enzymes; CBM, carbohydrate binding molecule; CE, carbohydrate esterase; CPIs, community phenotype indices; FAD, flavin adenine dinucleotide; FMN, flavin mononucleotide; GH, glycoside hydrolase; GT, glycosyltransferase; MCC, microcrystalline cellulose; NAD(P), nicotinamide adenine dinucleotide phosphate; RS4, resistant starch type IV; SLH, S-layer homology; TPP, thiamin pyrophosphate.

We then applied a subsystem-based approach using microbial community-centered implementation of the SEED database (mcSEED) to predict the metabolic potential of MAGs and metabolic phenotypes of the wider community (community phenotype indices [CPIs]; see Methods). Both fermentable fibres increased CPIs of succinate fermentation, serine biosynthesis, sorbitol and nucleoside utilization, while reducing CPIs of fructose utilization, cysteine biosynthesis, and transformation of cholic acid to 3-dehydrocholic acid and deoxycholic acid precursors (**Fig. S3e**, Supplementary Table 1). RS4 increased gamma-aminobutyric acid (GABA) production pathways, while AG increased acetate and indole-lactate production pathways, as well as pathways for utilization of various oligosaccharides (**Fig. S3e**, Supplementary Table 1).

### Fibre-specific effects on fecal SCFAs and bile acids

The fermentable fibres, but not MCC, altered fecal SCFA output (**Fig. 2c**). Both AG and RS4 increased total SCFAs concentrations (FDR-adjusted *p*=0.009 and *p*=0.027, respectively; linear mixed models). Interestingly, the two fibres differentially affected individual SCFAs: AG increased acetate (FDR-adjusted *p=*0.009) and propionate (FDR-adjusted *p*=0.008) while RS4 increased butyrate (FDR-adjusted *p*=0.027). In agreement with the overall increase in SCFAs, both fermentable fibres reduced fecal pH (FDR-adjusted *p*=0.023 and *p*=0.013 for AG and RS4, respectively) and the ratio of BCFAs to SCFAs (FDR-adjusted *p*=0.009 and *p*=0.013 for AG and RS4, respectively), indicative of increased fibre fermentation at the expense of proteolytic fermentation. Finally, while MCC did not affect SCFA levels, it significantly increased fecal dry mass percentage (FDR-adjusted *p*=0.008), which we speculate may be due to the 25g/35g dose of the non-fermentable fibre that is likely, to a large degree, excreted.

Given that fibre can bind bile acids, which affects their reabsorption and effects on human metabolism, and modulate the enzymatic capacity of the gut microbiome to transform bile acids into a large range of derivatives that are known to act on several receptors (*e.g.,* farnesoid X receptor, G protein-coupled bile acid receptor-1) to elicit anti-inflammatory effects ^46^, we applied targeted metabolomics to assess changes to fecal bile acid derivatives. We observed highly fibre-specific effects, with AG decreasing total fecal bile acids and eleven secondary bile acids, including dehydrolithocholic, 12-ketolithocholic, norursodeoxycholic, lithocholic, deoxycholic, dioxolithocholic, 12-ketochenodeoxycholic acids (all FDR-adjusted *p*<0.05; paired Wilcoxon tests) (**Fig. 2d**). Isolithocholic and alloisolithocholic acids were reduced by both AG and MCC, and MCC further reduced allocholic-3-sulfate and 7aOH-3-oxo-4-cholestenoic acid (all FDR-adjusted *p*<0.05). Notably, RS4 had no detectable effects on bile acid profiles (**Fig. 2d**).

### Characterization of the effect of fibre on mechanistic markers of host-microbiome interactions

We next assessed mechanistic markers of processes that are hypothesized to underpin host-microbiome interactions relevant for the immunometabolic benefits of fibre^8^. Specifically, we measured markers of gut barrier function (*e.g.,* fasting plasma lipopolysaccharide binding protein [LBP], fecal zonulin), intestinal and systemic inflammation (*e.g.,* fecal calprotectin, fasting plasma cytokines), and metabolic hormones [*e.g.,* fasting plasma ghrelin, PYY, leptin, GLP-1].

Plasma LBP was reduced by all three fibre: -11.5% by AG (FDR-adjusted *p*=0.001), -8.9% by RS4 (FDR-adjusted *p*=0.005), and -8.2% by MCC (FDR-adjusted *p*=0.012) (**Fig. 3a**). AG supplementation reduced fecal calprotectin, a marker of gut inflammation, by -33.8% (FDR-adjusted *p*=0.002) (**Fig. 3b**), with similar effect sizes also detected in the MCC group (-34.4%; FDR-adjusted *p*=0.017, linear mixed model). Systemic anti-inflammatory effects were also observed, but only for MCC, which reduced tumor necrosis factor-alpha (TNF-α) (FDR-adjusted *p*=0.011, ANCOVA models; **Fig. S4a**) and interleukin-6 (IL-6) (FDR-adjusted *p*=0.015; **Fig. S4b**) compared to RS4.

**Fig. 3.**
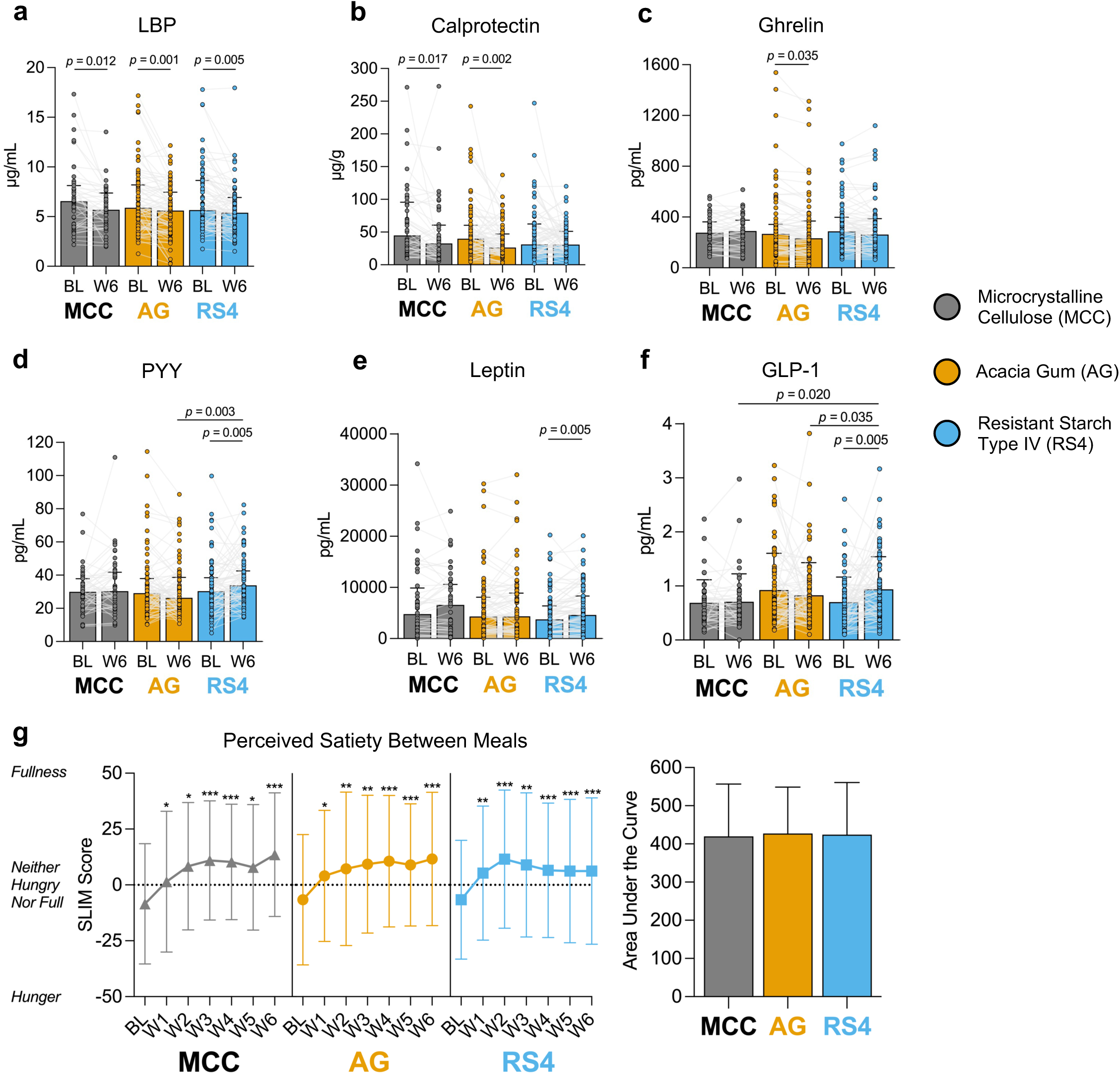
Effects of fibre supplements on mechanistic markers of host-microbiome interactions and perceived satiety. Bar plots show changes to mechanistic markers (**a-f**) that were significantly altered by fibre supplementation, with symbols representing individual participant changes. Data are presented as median ± IQR. FDR-adjusted *p* < 0.05 considered significant (within-group analyses – linear mixed models; between-group analyses – ANCOVA models). **g,** Changes in perceived satiety between meals assessed via the SLIM questionnaire. Asterisks represent within-group analyses (linear mixed models; * = *p* < 0.05, ** = *p* < 0.01, *** = *p* < 0.001). Area under the curve was calculated to assess differences between groups. Data are presented as mean ± SD. AG, acacia gum; BL, baseline; GLP-1, glucagon-like peptide 1; LBP, lipopolysaccharide binding protein; MCC, microcrystalline cellulose; PYY, peptide tyrosine tyrosine; RS4, resistant starch type IV; SLIM, Satiety Labeled Intensity Magnitude; W6, week 6.

In contrast to these universal effects, effects on circulating metabolic hormones that influence appetite and body weight were highly fibre-specific and only detected in the fermentable fibre groups. Compared to baseline, AG reduced ghrelin, an orexigenic (appetite-stimulating) hormone, by -7.2% (FDR-adjusted *p*=0.035) (**Fig. 3c**), whereas RS4 increased PYY, an anorexigenic (appetite-suppressing) hormone, by +10.8% (FDR-adjusted *p*=0.005) (**Fig. 3d**). RS4 also increased leptin, a hormone that decreases food intake and may increase energy expenditure, by +16.9% (FDR-adjusted *p*=0.005) (**Fig. 3e**). Finally, RS4 also increased GLP-1, an incretin hormone that stimulates insulin secretion and suppresses appetite, by +27.6% compared to baseline (FDR-adjusted *p*=0.005) and MCC (FDR-adjusted *p*=0.020, ANCOVA models) (**Fig. 3f**).

Other mechanistic markers of host-microbiome interactions (*e.g.,* serum trimethylamine-*N*-oxide [TMAO], fecal zonulin, and plasma IL-8, IL-10, adiponectin, and glucagon) were unaffected by the fibre supplements (**Fig. S4c-h**).

**Fig. S4.**
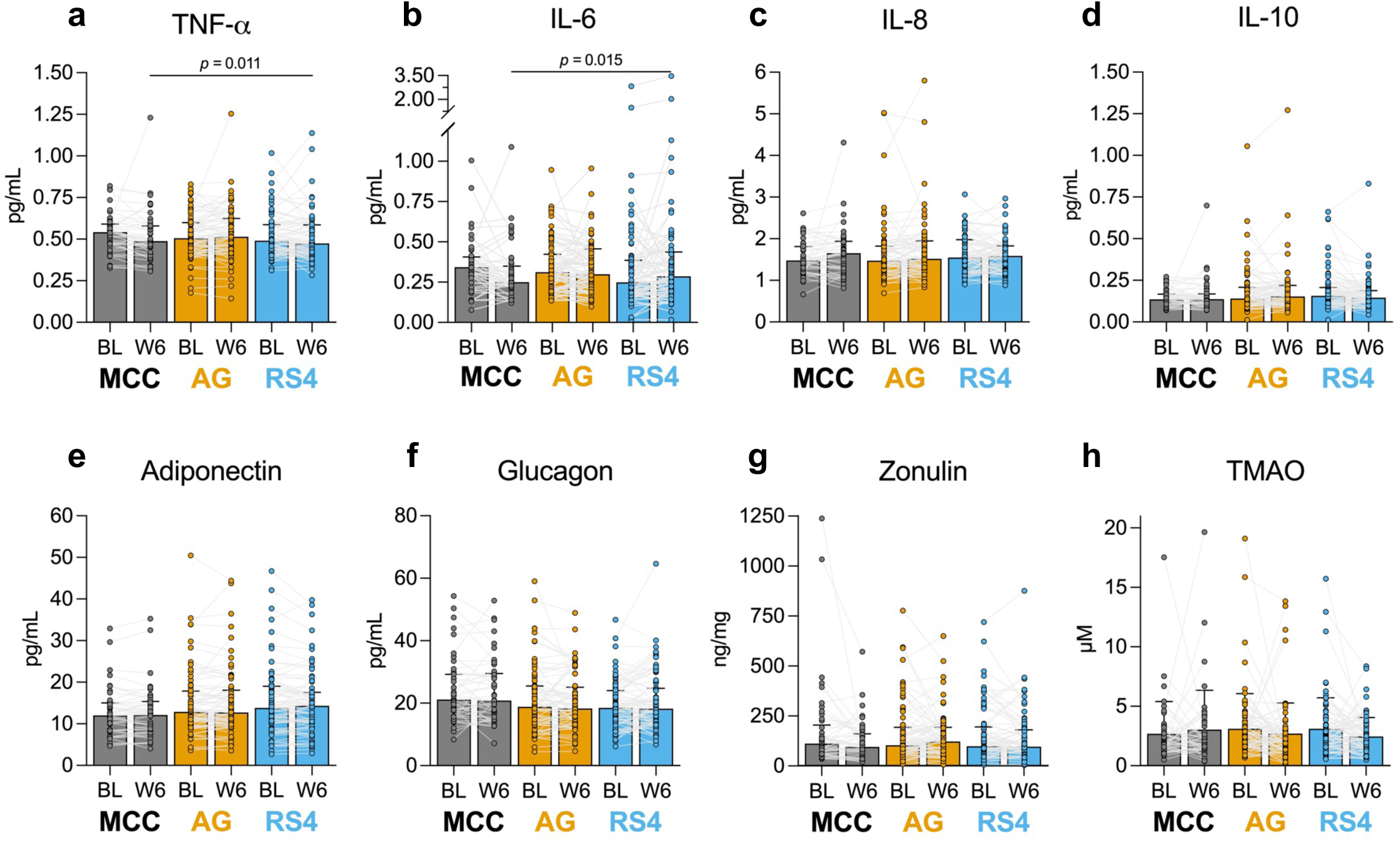
Effects of fibre supplements on hypothesized mechanistic markers of host-microbiome interactions. Bar plots show changes to mechanistic markers (**a-h**), with symbols representing individual participant changes. Data are presented as median ± IQR. FDR-adjusted *p* < 0.05 considered significant (within-group analyses – linear mixed models; between-group analyses – ANCOVA models). AG, acacia gum; BL, baseline; IL, interleukin; MCC, microcrystalline cellulose; RS4, resistant starch type IV; TMAO, trimethylamine-*N*-oxide; TNF-ɑ, tumor necrosis factor-alpha; W6, week 6.

### Fibre supplementation increase between-meal perceived satiety, but do not affect energy intake

We determined the effects of fibre supplementation on perceived satiety, assessed weekly during the intervention using the validated Satiety Labeled Intensity Magnitude (SLIM) questionnaire ^47^. All three fibres increased perceived satiety between meals over the entire treatment period (FDR-adjusted *p*<0.05 at all timepoints compared to baseline; linear mixed models), with participants on average feeling “slightly full” while consuming the fibres compared to feeling “slightly hungry” at baseline (**Fig. 3g**). Despite fibre-specific effects on satiety-related hormones, we saw no between-group differences in perceived satiety 30-60 mins before eating a meal, 30-60 mins and 2-2.5 hours after a meal, and before eating again (*i.e.,* between meals) (FDR-adjusted *p*>0.5; AUC of satiety measurements from baseline through to week 6, ANCOVA models).

Although fibre supplementation increased perceived satiety between meals, there were no concomitant reductions to energy intake captured by 24-hour dietary recalls (**Fig. S5a**). Protein intake increased significantly in the AG group from an average of 87.5 g/d to 103.4 g/d (FDR-adjusted *p*<0.001, linear mixed models) (**Fig. S5b**), corresponding to a change from 1.05 g/kg/day to 1.24 g/kg/day. Participants likely incorporated AG into protein-rich foods (*e.g.,* scrambled eggs, yogurt), though it is unclear why these changes were detected only in the AG group since all participants were given the same dietary instructions. Changes in protein intake did not influence perceived satiety, as including it as a covariate in statistical analyses did not alter results, nor was protein intake correlated with changes to ghrelin, LBP, or calprotectin (data not shown).

**Fig. S5.**
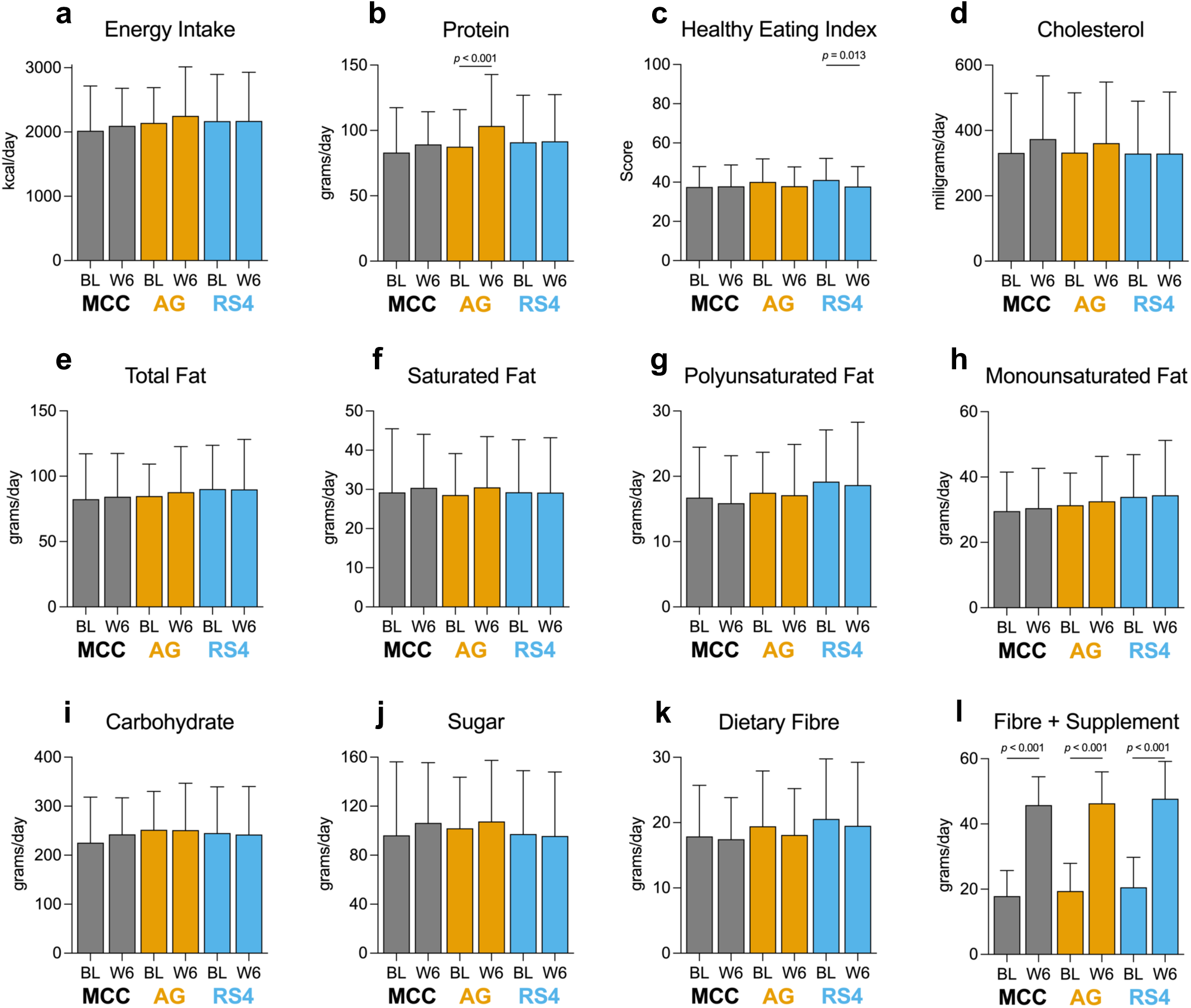
Changes to dietary intake during the fibre intervention. Dietary intake was assessed by 24-hour recalls, which captured changes to **a**, energy intake and **b-k,** macronutrients. **c**, Healthy Eating Index was calculated based on a food frequency questionnaire using the method outlined in Jessri et al. adapted for Canadian dietary guidelines. **l**, Total dietary fibre intake including amount of supplemented fibre at Week 6 (25g for females, 35g for males). Data are presented as mean ± SD. FDR-adjusted *p* < 0.05 considered significant (within-group analyses – linear mixed models). AG, acacia gum; BL, baseline; MCC, microcrystalline cellulose; RS4, resistant starch type IV; W6, week 6.

In the RS4 group, there was a small decrease in the healthy eating index (HEI), from an average of 41.0 to 37.8 (FDR-adjusted *p*=0.013; **Fig. S5c**). There were no clear displacements in any food groups that contributed to this change in HEI (data not shown), which is unlikely to be clinically relevant ^48^. There were no other group changes in dietary intake observed (**Fig. S5d-k**) except for fibre intake when accounting for the daily dose of the supplement, which significantly increased in all groups to over 45 g/d (**Fig. S5l**).

### No significant improvements in cardiometabolic risk markers with isolated fibre supplements at the group level

We assessed a comprehensive panel of cardiometabolic risk markers, including those related to hypertension, dyslipidemia, dysglycemia, and systemic inflammation. The fibre supplements, however, did not improve any of these risk markers, and there was considerable inter-individual variation in responses (**Fig. 4a-e**).

**Fig. 4.**
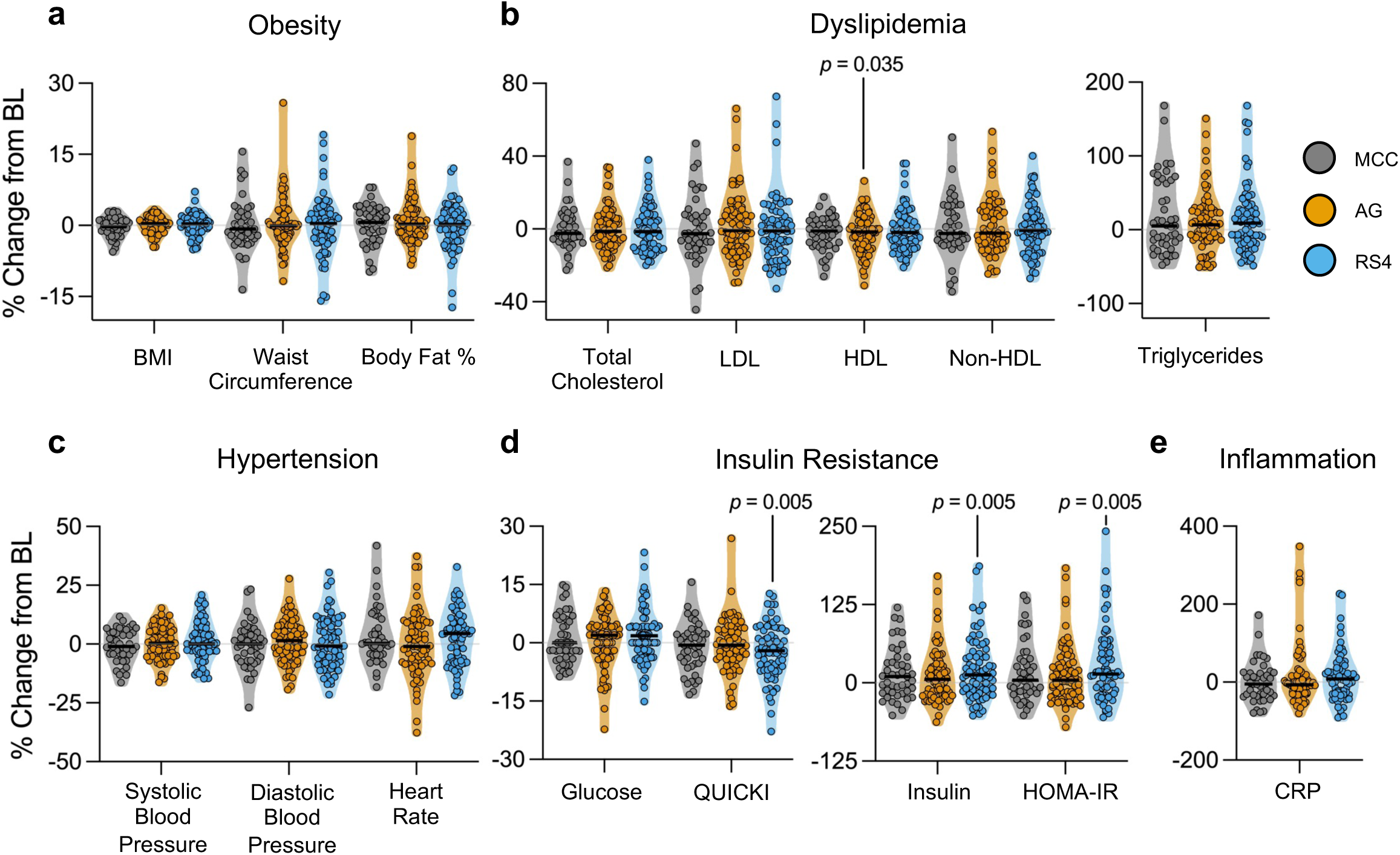
Inter-individual variation in responses to fibre supplementation for risk markers of chronic diseases. Violin plots show percent changes from baseline to week 6 in risk markers related to **a)** obesity, **b)** dyslipidemia, **c)** hypertension, **d)** insulin resistance, and **e)** systemic inflammation. Black lines represent medians, dots represent individual participants. FDR-adjusted *p* < 0.05 considered significant (within-group analyses – linear mixed models). AG, acacia gum; BL, baseline; BMI, body mass index; CRP, C-reactive protein; HDL, high-density lipoprotein cholesterol; HOMA-IR, homeostatic model assessment of insulin resistance; LDL, low-density lipoprotein cholesterol; MCC, microcrystalline cellulose; QUICKI, quantitative insulin-sensitivity check index; RS4, resistant starch type IV.

We detected two clinical effects of the fibre supplements that may be considered detrimental. AG decreased high-density lipoprotein (HDL) cholesterol by -1.7% (FDR-adjusted *p*=0.035, linear mixed model) (**Fig. 4b**). However, this change was small and likely not clinically relevant. RS4 increased fasting insulin by +12.7% (FDR-adjusted *p*=0.005), which drove increases in homeostatic model assessment of insulin resistance (HOMA-IR) of +14.6% (FDR-adjusted *p*=0.005) and a reduction in quantitative insulin-sensitivity check index (QUICKI) of -2.0% (FDR-adjusted *p*=0.005) (**Fig. 4d**). These effects of RS4 remained when changes in GLP-1 and HEI, which were altered in this group, were included as additional covariates in analyses (data not shown). We are unaware of other intervention or mechanistic studies showing similar effects of RS4 on insulin metabolism and, therefore, lack explanations for these effects.

### Analysis of the mechanistic underpinnings of the physiological effects of fibre supplementation

To gain mechanistic insight into the effects of fermentable dietary fibres, we determined if physiological effects (mechanistic markers of host-microbiome interactions and risk markers) could be predicted through fibre-induced changes in microbiome composition, functional features, diversity metrics, SCFAs, or bile acids. We developed individual machine learned models to predict mechanistic markers (calprotectin, LBP, ghrelin, PYY, GLP-1, and leptin) and representative risk markers (*i.e.,* those with highest correlation coefficients amongst other related variables; **Fig. S6a**) of obesity (BMI), hypertension (diastolic blood pressure), dysglycemia (HOMA-IR), dyslipidemia (total cholesterol), and systemic inflammation (C-reactive protein, CRP). For the risk markers, participants were classified as either ‘responders’ or ‘non-responders’ based on whether they were in the bottom (*i.e.,* reduction in the marker) or top (*i.e.,* increase in the marker) tertile, respectively (**Fig. S6b**). For mechanistic markers, ‘responders’ and ‘non-responders’ were instead defined based on the cohort median. Using five-times repeated 10-fold cross-validation, we developed independent random forest classification models (**Fig. S6c**) trained on microbiome features separately (composition [MAGs, species, ASVs], functions [pathways, CAZymes, aggregated abundances of CAZymes, CPIs, SCFAs, bile acids], or diversity metrics). We also combined these individual datasets (*i.e.,* all features combined) to determine if model performance would be augmented, as previously done in other studies ^49,50^. We only consider models with cross-validated accuracies ≥70% and area under the receiver operating characteristic curves (AUROC) ≥0.70 to be of sufficient accuracy.

**Fig. S6.**
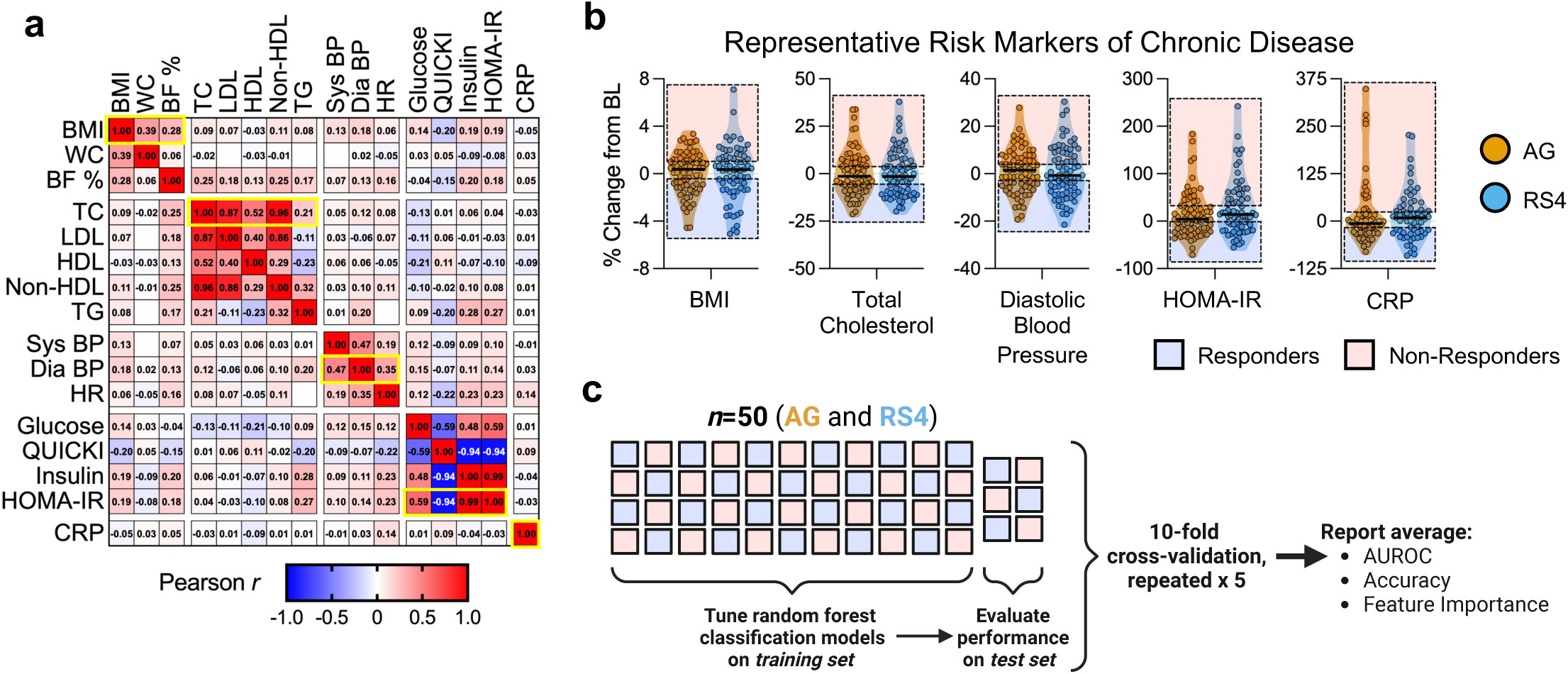
Machine learning strategy to predict responses to fibre supplementation. **a**, Correlation matrix of associations between risk markers of chronic diseases, with Pearson coefficients shown in each box (boxes that are empty have *r* < 0.01). Yellow boxes highlight the risk markers within each group that had the highest correlation coefficients with other markers in its class (‘representative risk markers of chronic diseases’). **b**, Violin plots show percent changes from baseline to week 6 in representative risk markers, and highlight the classification of participants (dots) as either responders (reduction in marker; blue box) or non-responder (increase in marker; red box). **c**, Schematic of machine learning approach. Figure created in Biorender. AG, acacia gum; AUROC, area under the receiving operator characteristic curve; BL, baseline; BMI, body mass index; BF %, body fat percentage; BP, blood pressure; CRP, C-reactive protein; HDL, high-density lipoprotein cholesterol; HOMA-IR, homeostatic model assessment of insulin resistance; HR, heart rate; LDL, low-density lipoprotein cholesterol; MCC, microcrystalline cellulose; QUICKI, quantitative insulin-sensitivity check index; TC, total cholesterol; TG, triglycerides; RS4, resistant starch type IV.

In the AG group, ghrelin responses were predicted by shifts in aggregated abundances of CAZymes (accuracy 70%, AUROC=0.76) (**Fig. 5a** and Supplementary Table 2). Interestingly, top predictive features (*i.e.,* those with the highest average importance scores) were enzymes that target the carbohydrate moieties of AG, such as aggregated abundances of CAZymes targeting arabinogalactan and galactan, as well as arabinoxylan and xylan from similar CAZyme families ^51^, of which responders had greater increases on average (**Fig. 5b**). Ghrelin concentrations were negatively associated with aggregated abundances of CAZymes that target arabinogalactan (*rs* = -0.2), arabinan (*rs* = -0.29), and galactan (*rs* = -0.28) (all FDR-adjusted *p=*0.09; **Fig. S7a**), suggesting that reductions in ghrelin are linked to increased fermentative capacity towards AG.

**Fig. 5.**
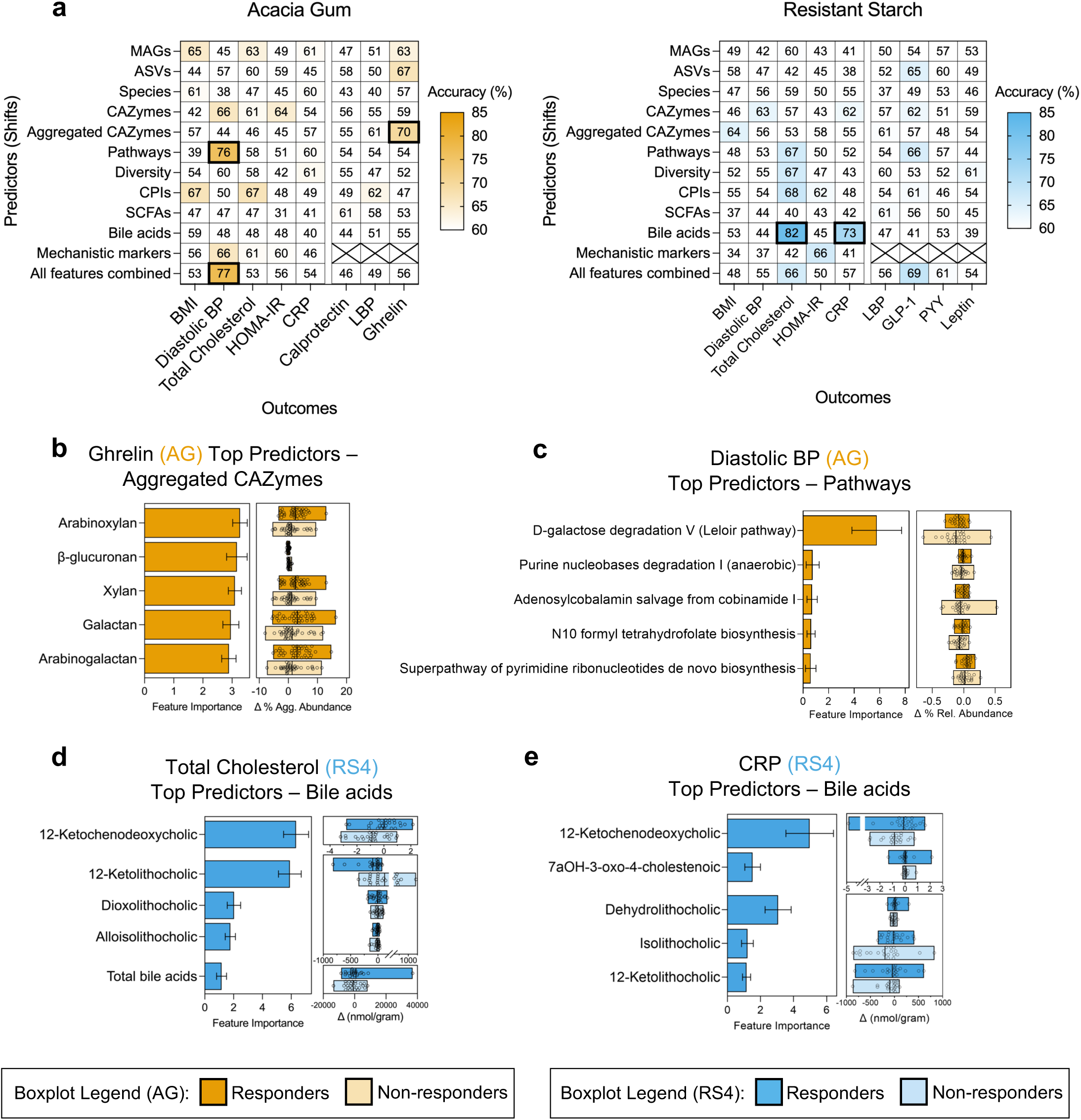
Prediction of fibre responses based on shifts in features significantly altered by fibre supplementation to gain potential mechanistic insight. Random forest classification models were used to predict responses in representative risk markers of chronic diseases as well as mechanistic markers that were altered by fibre supplementation in the AG and RS4 groups separately. **a,** Heatmap showing cross-validated accuracy values for individual datasets. Models that had cross-validated accuracy >70% and AUROC >0.70 are outlined with a black box, and their top predictive features are shown in: **b,** aggregated CAZyme abundances to predict ghrelin responses in the AG group; **c,** microbial pathways to predict diastolic blood pressure responses in the AG group; **d,** fecal bile acids to predict total cholesterol responses in the RS4 group; **e,** fecal bile acids to predict CRP responses in the RS4 group. Features are visualized as bar plots with mean ± SD feature importance, and boxplots show differences between responders (dark colour) and non-responders (light colour); dots are individual participants. AG, acacia gum; ASV, amplicon sequence variants; AUROC, area under the receiving operator characteristic curve; BMI, body mass index; CAZymes, carbohydrate-active enzymes; CPIs, community phenotype indices; CRP, C-reactive protein; GLP-1, glucagon-like peptide 1; LBP, lipopolysaccharide binding protein; MAGs, metagenome assembled genomes; PYY, peptide tyrosine tyrosine; RS4, resistant starch type IV; SCFAs, short-chain fatty acids.

**Fig. S7.**
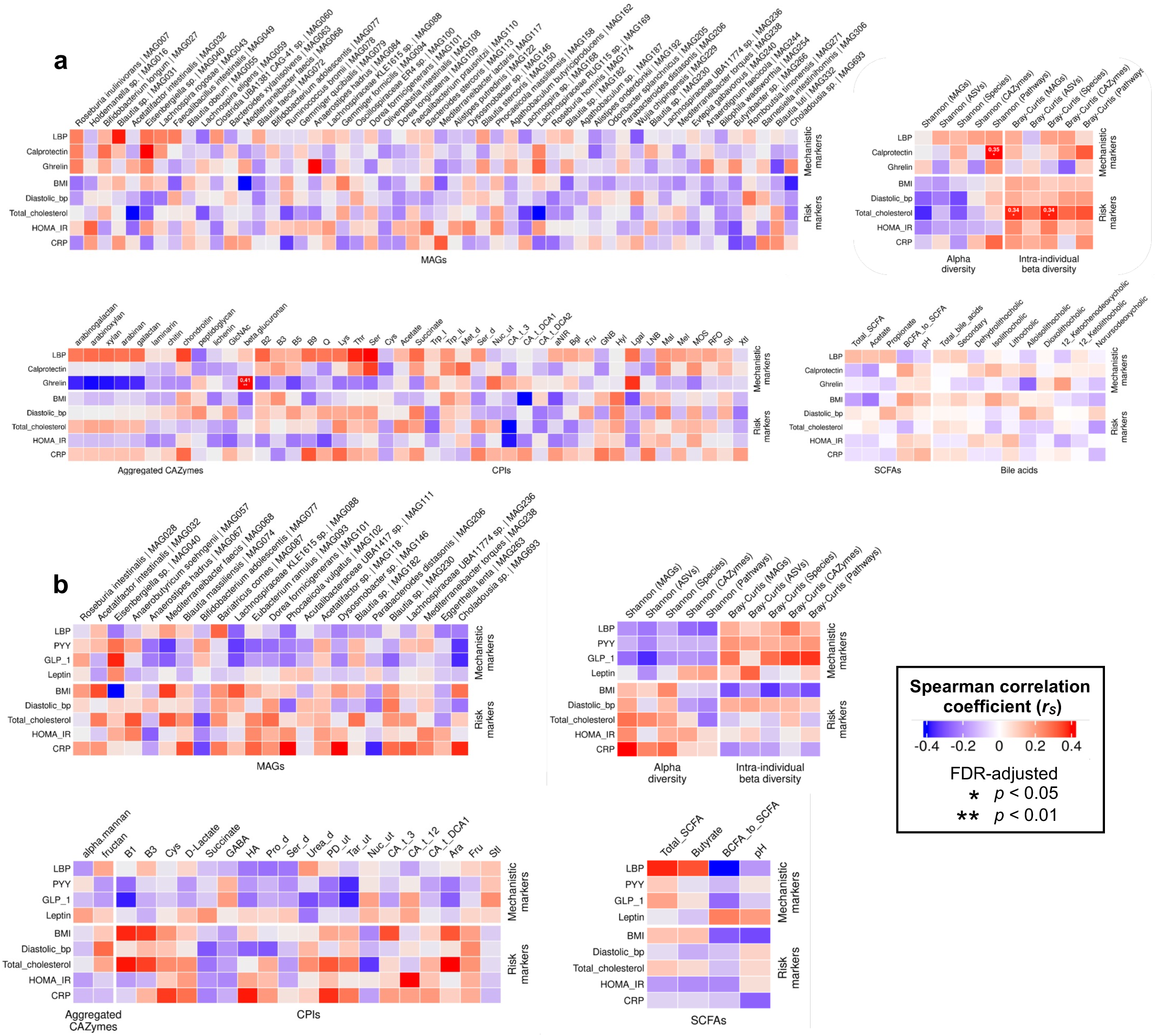
Links between fibre-specific shifts in microbiome features with health outcomes. Heatmaps showing Spearman correlation coefficients (*r_s_*) in the (**a**) AG and (**b**) RS4 groups between shifts [log_10_(W6/BL)] in mechanistic markers of host-microbiome interactions/risk markers of chronic diseases and MAGs; alpha diversity (Shannon Index) and intra-individual beta diversity (Bray-Curtis dissimilarity between baseline and week 6 samples of the same participant, not log-transformed); aggregated CAZymes and CPIs; SCFAs and bile acids. AG, acacia gum; ASV, amplicon sequence variant; BCFA, branched-chain fatty acid; BMI, body mass index; BP, blood pressure; CAZymes, carbohydrate-active enzymes; CPIs, community phenotype indices, CRP, C-reactive protein; GLP-1, glucagon-like peptide 1; HOMA-IR, homeostatic model assessment of insulin resistance; LBP, lipopolysaccharide binding protein; MAGs, metagenome assembled genomes; PYY, peptide tyrosine tyrosine; RS4, resistant starch type IV; SCFAs, short-chain fatty acids; TC, total cholesterol.

Diastolic blood pressure responses were predicted by shifts in microbial pathways (accuracy 76%, AUROC=0.75) (**Fig. 5a** and Supplementary Table 3). The feature with the highest importance, D-galactose degradation Y (Leloir pathway) (**Fig. 5c**), might contribute to AG utilization as D-galactose is a main constituent of arabinogalactan ^52^. Predictions were slightly augmented by the pooled feature set (accuracy 77%, AUROC=0.82) (**Fig. 5a** and Supplementary Table 3). Top predictive features again included the D-galactose degradation Y (Leloir pathway), as well as the CAZyme subfamily GH43_23 that can target arabinogalactan ^53^, features that were, on average, increased in responders. Taken together, inter-individual variation in diastolic blood pressure responses may be explained by differences in the degradation capacity of the microbiome towards the structure of AG.

Despite no significant changes to bile acids in the RS4 group (**Fig. 2d**), individual responses in both total cholesterol and CRP were predicted by shifts in fecal bile acids (**Fig. 5a** and Supplementary Table 3). For total cholesterol (accuracy 82%, AUROC=0.85), top predictive features included 12-ketolithocholic acid and dioxolithocholic acid, secondary bile acids that were, on average, reduced in responders, as well as 12-ketochenodeoxycholic acid that, on average, was reduced in non-responders (**Fig. 5d**). For CRP (accuracy 73%, AUROC=0.70), some top predictive features overlapped with those for total cholesterol (*i.e.,* 12-ketolithocholic acid and 12-ketochenodeoxycholic acid), but also included dehydrolithocholic and isolithocholic acids (**Fig. 5e**).

A surprising result of our mechanistic analysis was that neither the machine learning approach nor a correlation analysis did identify microbiome features considered classic mediators of prebiotic effects of fibre, such as selective shifts in bacterial taxa (*e.g., Bifidobacteria* species) or SCFAs, as predictors of the physiological effects of the fermentable fibres, even though these were altered by the interventions (**Fig. S7**).

### Clinical responses to fermentable fibres can be predicted using baseline microbiome signatures

Given that the gut microbiome ferments fibre structures into bioactive metabolites, while being remarkably stable but individualized, we hypothesized that baseline microbiome signatures could accurately predict clinical responses to the intervention. We applied the same machine learning approach as above but used baseline microbiome features, as well as host features such as baseline dietary intake and metabolic status, to predict responses in mechanistic markers and representative risk markers of chronic diseases.

In the AG group, diastolic blood pressure was best predicted by microbial pathways, with 72% accuracy (AUROC=0.77) (**Fig. 6a** and Supplementary Table 4). Top predictive pathways were those involved in carbohydrate metabolism (*e.g.,* mannan degradation, sucrose degradation), of which responders had higher levels at baseline (**Fig. 6b**). In the RS4 group, baseline fecal bile acid concentrations predicted changes to CRP, with 70% accuracy (AUROC=0.74) (**Fig. 6a** and Supplementary Table 4). Top predictive features were glycine-conjugated bile acids, of which responders had higher levels at baseline (**Fig. 6c**). Baseline microbiome signatures did not predict responses in mechanistic markers with high accuracy (<70% for all models; Supplementary Table 5). Furthermore, contrary to previous reports ^54–56^, baseline dietary intake, including habitual fibre consumption and diet quality, did not accurately predict any physiological responses to fibre (**Fig. 6a**, Supplementary Tables 4 and 5).

**Fig. 6.**
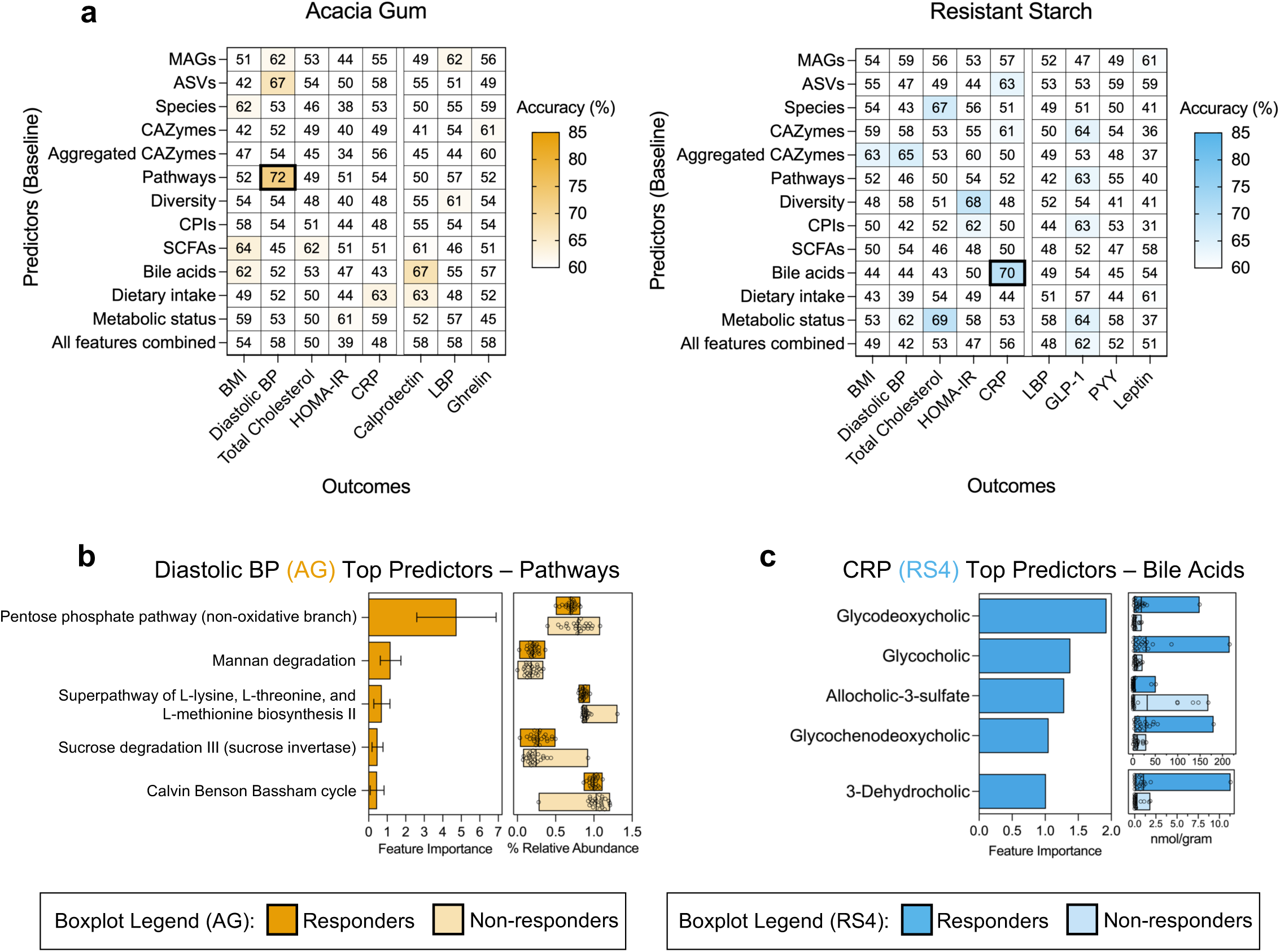
Prediction of fibre responses using on baseline features. Random forest classification models were used to predict responses in representative risk markers of chronic diseases as well as mechanistic markers that were altered by fibre supplementation in the AG and RS4 groups separately. **a,** Heatmap showing cross-validated accuracy values for individual datasets. Models that had cross-validated accuracy >70% and AUROC >0.70 are outlined with a black box, and their top predictive features are shown in: **b,** microbial pathways to predict diastolic blood pressure responses in the AG group, and **c,** fecal bile acids to predict CRP responses in the RS4 group. Features are visualized as bar plots with mean ± SD feature importance, and boxplots show differences between responders (dark yellow/blue) and non-responders (light yellow/blue); dots are individual participants. AG, acacia gum; ASV, amplicon sequence variants; AUROC, area under the receiving operator characteristic curve; BMI, body mass index; BP, blood pressure; CAZymes, carbohydrate-active enzymes; CPIs, community phenotype indices; CRP, C-reactive protein; GLP-1, glucagon-like peptide 1; LBP, lipopolysaccharide binding protein; MAGs, metagenome assembled genomes; PYY, peptide tyrosine tyrosine; RS4, resistant starch type IV; SCFAs, short-chain fatty acids.

We then pooled the individual datasets (baseline MAGs, CAZymes, aggregated abundances of CAZymes, pathways, diversity metrics, CPIs, SCFAs, bile acids, metabolic status, and dietary intake) to determine if combining baseline microbiome- and host-derived features would outperform the predictive performance of the individual datasets. However, for both AG and RS4, this approach did not improve predictive accuracy for any physiological responses compared to the highest-performing models based on individual datasets (**Fig. 6a**, Supplementary Tables 4 and 5).

## DISCUSSION

This study addressed critical questions about the beneficial health effects of isolated dietary fibres. Specifically, it examined the role of gut microbiome fermentation, the mechanisms linking fibre-microbiome interactions to host physiology, and the basis for inter-individual variability in clinical responses. Our findings indicate that while structurally-distinct fermentable fibres, AG and RS4, produce clear, fibre-specific changes in gut microbiome composition and SCFAs, these shifts show no detectable links to physiological benefits. Instead, effects on gut barrier function (LBP), gut inflammation (calprotectin), satiety, and systemic inflammation (TNF-α, Il-6) were either detectable in both the fermentable (AG and RS4) and non-fermentable fibre (MCC) groups, or only in MCC, suggesting fermentation-independent effects. In contrast, effects of AG on ghrelin and blood pressure, and RS4 on cholesterol and CRP, were linked to shifts in microbiome features involved in fibre utilization and signalling (bile acids). Moreover, effects on blood pressure (with AG) and CRP (with RS4) were predictable using baseline microbiome features. Collectively, these results point to both microbiome independent and dependent effects of fibre supplements, and provide evidence that inter-individual differences in microbiome functions contribute to variable clinical responses. Our findings can, thus, inform a mechanistic framework for the application of fibre supplements in a targeted and personalized manner.

The fermentable fibres, AG and RS4, caused highly selective microbiome changes, enriching for putative health-promoting *Bifidobacterium*, *Faecalibacterium,* and *Lachnospiraceae* species. These fibres also had discrete effects on individual SCFAs, confirming previous work that proposed that microbiome metabolism can be precisely directed by distinct fibre structures ^33,57^. Several metabolic hormones that are inducible through SCFAs via G-protein–coupled receptors on enteroendocrine cells were affected by AG (ghrelin) and RS4 (PYY, leptin, and GLP-1). However, these effects were neither predicted by nor correlated with the microbial taxa or SCFAs enriched by either AG or RS4. Our study, therefore, does not provide evidence that selective microbiome alterations induced by dietary fibres are directly relevant for health outcomes, a concept that is germane to the mode of action of prebiotics ^14^.

By comparing structurally-distinct fermentable (AG and RS4) and non-fermentable (MCC) fibres, our study design allowed for causal inferences to be made on whether physiological effects of fibre are driven by microbial fermentation. Some of the strongest effect sizes observed were reductions in fecal calprotectin and plasma LBP by AG, yet MCC also reduced both markers by a similar magnitude and further showed systemic anti-inflammatory effects. In addition, improvements of these markers could not be predicted by baseline or shifts in microbiome features. Overall, our results indicate that these effects were fermentation independent. The underlying mechanisms remain unclear, but non-fermentable cellulose has been shown to alter transcriptional responses in colonic epithelial cells, reducing intestinal apoptosis and inflammation in animal models of colitis while enhancing gut barrier function ^58,59^. MCC and AG further selectively altered fecal bile acid derivatives, reducing secondary bile acids that may be cytotoxic and pro-inflammatory ^60^. Experiments in germ-free mice demonstrated that the metabolic and immunological effects of fibres (RS) can be completely microbiome independent, with improvements in insulin sensitivity mechanistically linked to altered bile acid profiles and adipose tissue immune modulation ^61^. Future research is needed to elucidate the exact mechanisms by which non-fermentable fibres improve host physiology in humans.

The three-arm comparison of two fermentable fibres with MCC also revealed effects that are likely microbiome dependent. Several outcomes were specific to a single fermentable fibre and could be mechanistically linked to microbial functions. Reductions in fasting ghrelin with AG were predicted by, and correlated with, increases in the community-level abundance of CAZymes targeting AG structural components (*e.g.,* arabinogalactan, galactan). Similarly, diastolic blood pressure responses to AG were associated with shifts in the Leloir pathway for D-galactose degradation and enrichment of the CAZyme subfamily GH43_23 (containing α-L-arabinofuranosidases, which hydrolyze terminal α-L-arabinofuranose residues from arabinogalactans), both features likely contributing to AG utilization ^52,53^. Together, these findings suggest that the physiological effects of AG on ghrelin and blood pressure are shaped by the functional capacity of the microbiome to degrade its specific fibre structure. These associations are consistent with human evidence that both microbial fibre fermentation and SCFAs can suppress postprandial ghrelin^62^ and lower blood pressure^63,64^.

Although we did not detect clear structure-function relationships in the RS4 group, fibre-induced shifts in fecal secondary bile acids predicted personalized responses in total cholesterol and CRP. A variety of gut microbes deconjugate bile acids ^65^, thereby promoting hypocholesterolemic effects ^66^, while bile acids also act as signalling molecules that regulate immune homeostasis ^67^. These findings suggest that RS4 modifies microbially derived bile acid metabolism in an individualized manner, with potential health effects beyond traditional prebiotic endpoints.

The primary objective of this study was to determine the ability of the gut microbiome at baseline to predict clinical improvements of the dietary fibre treatments. Only two machine learned models performed with acceptable quality, one for AG and one for RS4. The effects of AG on blood pressure were predicted by baseline microbiome pathways, with the non-oxidative branch of pentose phosphate pathway being the top predictor. This pathway may be central to AG utilization by gut microbes, as it channels arabinose released from its arabinogalactan structure into pentose phosphates that are converted into glycolytic intermediates that can be used to produce essential building blocks, such as nucleotides and amino acids ^68^. Greater baseline abundance of this pathway could, therefore, favour assimilation of arabinose-derived carbon into microbial biomass rather than fermentation end-products, attenuating blood pressure responses. Good predictions were also obtained for CRP responses in the RS4 group, with baseline fecal bile acids as the best predictors. Together, these results provide proof-of-concept that baseline microbiome features can explain high inter-individual variation in certain clinical responses, thereby helping to identify individuals who would most likely benefit from specific fibres, and that the predictors align with plausible microbial mechanisms. Such microbiome-informed stratification could enable targeted fibre interventions with measurable clinical impact, an approach particularly relevant here, as neither fibre produced significant benefits at the cohort level.

The lack of consistent, population-wide effects of the isolated fibre supplements on risk markers of chronic diseases is notable, especially compared to the consistent effects of high-fibre, whole-food approaches on exactly these markers ^23,69–71^. This discrepancy is increasingly acknowledged in the nutrition literature, raising questions about the clinical efficacy of fibre supplements relative to the intrinsic fibres naturally present in whole-plant foods ^1,28^. Intrinsic fibres are embedded in complex cell-wall matrices and bound to diverse bioactive compounds that provide health effects not preserved in isolated fibre types ^72^. In addition, the structural and chemical context in which intrinsic fibres exist reduces nutrient accessibility and protects them from rapid fermentation, supporting more gradual and distal SCFA delivery ^28,73^. In contrast, single isolated fibres, like the ones used in our study, lack this complexity and might be fermented in more proximal regions of the gut, with the exact location of fermentation potentially differing among individuals depending on their microbiome. This is important as consistent immunometabolic effects of dietary fibre have been shown with only distal SCFA delivery, not proximal ^74,75^, leading to the hypothesis that distal colonic saccharolytic fermentation is a key modulator of factor preventing chronic metabolic diseases ^76^.

Although isolated fibres lack several of the key functional characteristics of intrinsic fibres, the findings of our study suggest that inconsistent clinical benefits do not reflect a lack of efficacy per se. Instead, effects are specific to particular targets, which were, in our case, mechanistic markers linked to inflammation, gut barrier function, and gut hormones. In a different clinical context, such effects could be highly beneficial. The effects on cardiometabolic risk markers were highly individualized and predictable through microbiome functional capacity or fibre-specific interactions with bile acid pools. Importantly, we demonstrate that mechanistic features of fibre utilization and bile acid metabolism can be identified at baseline and predict clinical responses, providing a basis for the personalized use of fibre supplements. Finally, we provide evidence for fermentation-independent effects of both fermentable and non-fermentable fibres, pointing to underexplored pathways that could be systematically leveraged in future applications.

Such knowledge provides key mechanistic insight into the drivers of individualized clinical effects of fibre supplements ^25,77^, and it enables several complementary strategies to overcome them. It supports a more systematic application of fibre structures to defined mechanistic targets (microbiome-dependent or independent), tailoring fibre use to an individual’s microbiome configuration and designing fibre mixtures that broaden microbial interactions and mitigate individualized responses. Overall, our work provides a roadmap for moving beyond generic supplementation strategies towards mechanistically guided, personalized interventions that consider fibre structure, site of action in the gut, and inter-individual variability. Such an approach can make fibre supplementation more consistently effective and clinically meaningful within precision nutrition frameworks.

### Limitations of study

Despite several strengths of this intervention study, there are limitations. The human trial was conducted in participants that were advised to maintain their habitual diet. However, since there are only certain foods/beverages the fibres can be mixed into successfully, supplementation can cause other foods and nutrients to be displaced. This was evident in the AG group, which on average increased daily protein intake during the intervention likely due to the foods/beverages that were used as vehicles for the fibre. Fecal samples do not accurately capture *in vivo* SCFA production, which may have led to them not being detected as predictors of the physiological effects of the fermentable fibre even though genetic features contributing to fibre utilization were. Future intervention studies may consider employing alternative methods, such as capsule devices ^78,79^, to capture SCFA dynamics *in vivo.* While the total sample size of *n*=195 exceeds that of most fibre intervention studies ^25,27^, the machine learning applied within each arm likely would have been strengthened with a much larger sample size and dataset for the predictive models ^80,81^. Further, the lack of an independent test set risks overfitting of the models to the cohort-specific data; however, external cross-validation for model evaluation was applied to help partially overcome this potential bias.

## METHODS

### Study design and ethical procedures

The Alberta FYBER Study (Feed Your gut Bacteria morE fibeR) was a single-center, six-week, parallel, three-arm, single-blinded, randomized controlled trial (**Fig. 1a**) conducted at the Human Nutrition Research Unit at the University of Alberta (Edmonton, AB, Canada) in accordance with the principles of the Declaration of Helsinki. This trial was prospectively registered on 3 July 2015 on ClinicalTrials.gov (identifier NCT02322112; https://clinicaltrials.gov/study/NCT02322112), and initially included an additional fibre intervention arm of arabinoxylan. However, in response to grant reviewer comments that advised against including this premarket fibre in a large human trial, the arabinoxylan arm was removed on October 26^th^, 2016 and the 15 participants who had already completed the protocol were analyzed separately ^42,43,82^. The protocol was approved by the University of Alberta’s Health Research Ethics Board (Pro00050274). Study data was collected and securely managed using REDCap electronic data capture tools hosted and supported by the Women and Children’s Health Research Institute at the University of Alberta ^83^.

### Subjects

Healthy men and pre-menopausal, non-pregnant, and non-lactating women, aged 19-50 years, with a BMI of 25-35 kg/m^2^ were recruited using online advertisements and flyers. Only participants who provided written consent were enrolled in the study. We chose the BMI range of 25-35 kg/m^2^ as it has been linked to elevated levels of risk markers of chronic diseases, especially systemic inflammation (*i.e.,* CRP) that increases risk for obesity-related pathologies ^84,85^. Exclusion criteria were: (1) history of gastrointestinal disorders or surgeries; (2) history of diabetes mellitus; (3) chronic use of anti-inflammatory, anti-diabetic, anti-hypertensive, lipid-lowering, or laxative medications; (4) alcohol intake ≥7 drinks/week; (5) >3 hours of moderate-vigorous exercise per week; (6) allergy or intolerance to treatment fibres; (7) vegetarian or vegan; (8) smoking; (9) antibiotic use three months prior to enrollment; (10) use of probiotic, prebiotic, omega-3 fatty acid, or herbal supplements. If subjects used any of these supplements but were otherwise eligible, they could elect to undergo a four-week washout period before participating in the study.

Since machine learning models based on microbiome features that predict effects of dietary fibre have not yet been established, it was difficult to power our study. We aimed to enroll *n* = 75 in each of the fermentable fibre arms and *n* = 50 in the control arm, which exceeds subject numbers used in studies that successfully established an effect of fibre supplementation on inflammation, glucose metabolism, and lipid metabolism in healthy overweight individuals ^25^. However, given the slower-than-expected recruitment of study participants, and that predictive models would not be developed based on the gut microbiome for the non-fermentable control fibre, the MCC arm was capped at *n* = 45 and recruitment was then prioritized for the AG and RS4 arms.

### Randomization and blinding

Random treatment allocation was conducted using a computerized random number generator, as previously described ^42^. Subjects were stratified based on sex and randomly assigned to one of the three study arms by a researcher not involved in data analysis. If a participant withdrew from the study, an additional subject was enrolled to replace them using the same random allocation methods.

The fibre supplements were packaged in opaque paper bags to blind participants to fibre assignment. Investigators were not blinded as the fibres were not identical in appearance, and the investigators provided participants with fibre-specific suggestions for incorporation into foods.

### Experimental protocol

Participants were provided individual daily sachets of their assigned fibre to incorporate into their usual diet for six weeks. To align with recommendations for dietary fibre intake ^86^, participants consumed 25 g/d (females) or 35 g/d (males), with the exception of days 1 and 2 where half-doses (12.5 or 17.5 g/d for females and males, respectively) were provided to ease incorporation of the fibre into the diet. Daily doses were corrected for the purity of the three fibres (detailed below) so that all participants received 25 or 35 g/day of dietary fibre, regardless of treatment. Participants incorporated the powdered supplements into their preferred foods and drinks (*e.g.,* yogurt, smoothies, scrambled eggs, etc.), and were otherwise instructed to maintain their usual dietary habits throughout the intervention. This was assessed through administration of 24-hour dietary recalls (detailed below). Participants were asked to return all fibre sachets so that any remaining fibre could be weighed to assess adherence. If participants were <80% adherent, they were removed from the study.

### Dietary fibre supplements

The AG was Agri-Spray Acacia^®^ R (Agrigum^®^ International Limited, Buckinghamshire, UK), a fibre isolated from the exudate of the Acacia tree (*Acacia senegal*) that contained ∼85% dietary fibre. AG is primarily comprised of arabinogalactan, containing β-(1,3) and β-(1,6) galactose and α-(1,5) and α-(1,3) arabinose branches, is water-soluble, fermentable, and has low-viscosity ^40^. The RS4 was Fibersym^®^ RW (MGP^®^ Ingredients, KS, USA), a phosphorylated, cross-linked, wheat-based resistant starch that consisted of linear α-(1,4) and α-(1,6) glucose branches and contained ∼85% dietary fibre. RS is an insoluble yet fermentable fibre that displays properties similar to that of soluble fibres in the gastrointestinal tract ^41^. The MCC was Microcel^®^ MC-12 (Blanver Farmoquimica LTDA, São Paulo, Brazil), a large-particle (160-micron average), wood-derived cellulose deprived of hemicellulose and amorphous regions that consisted of linear β-(1,4) glucose branches and contained around 95% dietary fibre. MCC is water-insoluble, non-viscous, and previous work confirms it is non-fermentable by human gut microbiota as it had no effect on microbial compositional or functional features ^42^.

### Specimen collection

Participants were provided stool collection kits (consisting of a stool specimen container [Fisher, Canada], a GasPak™ EZ Anaerobe Sachet [BD, Canada] to generate an anaerobic environment, and an airtight plastic bag) to collect fecal samples at baseline and week 6 of the intervention. Fecal samples were received within four hours of collection and processed immediately in an anaerobic chamber (5% CO2, 5% H2, 90% N2). Aliquots of raw fecal material (for bile acid profiling), fecal material diluted 1:10 in sterile phosphate-buffered saline (PBS) (for DNA extraction), and fecal material diluted 1:5 in sterile 5% phosphoric acid (for SCFA and BCFA measurements) were frozen at - 80°C for downstream analyses (all performed after trial completion to avoid batch effects).

Blood samples were collected after an overnight fast (≥12 hours) via venipuncture at baseline and week 6 of the intervention using serum separation, K2-ethylenediaminetetraacetic acid- (EDTA) coated, and P800 BD Vacutainer^®^ collection tubes (Becton, Dickinson and Company, NJ, USA). P800 tubes were pre-coated with K2EDTA and a proprietary cocktail of protease, esterase, and DPP-IV inhibitors to prevent degradation of hormones. After centrifugation, serum, plasma, and inhibitor-treated plasma were aliquoted and stored at −80°C for downstream analyses. Whole blood was also analyzed for complete blood count (red blood cells, hemoglobin, hematocrit, platelets, and white blood cells), on a Sysmex XN-10™ Automated Hematology Analyzer (Sysmex, IL, USA).

### Outcome measurements

Gut microbiome features (diversity indices, compositional features, pathways, CAZymes, CPIs, SCFAs, and bile acids), mechanistic markers of host-microbiome interactions (calprotectin, LBP, zonulin, metabolic hormones, inflammatory cytokines, and trimethylamine-*N*-oxide [TMAO]), risk markers of chronic diseases (BMI, body fat percentage, waist circumference, blood pressure, glucose, insulin, lipid panel, and CRP), and dietary intake were assessed at baseline and week 6 of the intervention. To assess changes to gastrointestinal symptoms and perceived satiety throughout the intervention, questionnaires were administered at baseline and weekly throughout the intervention.

#### DNA extraction and microbiome sequencing

One mL of raw fecal material diluted in PBS (*i.e.*, 0.1 g of raw feces) was centrifuged at 8000 g for 5 minutes at 4°C to obtain fecal pellets, and microbial DNA was extracted from the fecal pellets using the QIAamp PowerFecal Pro DNA Kit (QIAGEN, Germany), following the manufacturer’s protocol. The V5-V6 hypervariable region of the 16S rRNA gene was amplified using the primer pair 784F (5′-RGGATTAGATACCC-3’) and 1064R (5′-CGACRRCCATGCANCACCT-3′), and the amplicons were paired-end sequenced (2*300 bp) using Illumina MiSeq PE300 at the University of Minnesota Genomics Center (MN, USA). Extracted DNA was also subjected to WMS. Metagenome libraries of each DNA sample were generated using the NEBNext Ultra II DNA Library Prep Kit for Illumina (New England Biolabs, ON, Canada). The libraries were sequenced using NovaSeq6000 S4 PE150 platform (2*150 bp paired-end sequencing) at the Génome Québec Innovation Centre (QC, Canada).

#### Fecal SCFAs, pH, dry mass percentage, and bile acids

Fecal SCFA quantification was performed using gas chromatography by the Agricultural, Food and Nutritional Science Chromatography Facility at the University of Alberta, as previously described ^42^. Total SCFA concentrations were determined as the sum of acetate, propionate, butyrate, and valerate, and the relative proportions of each were determined by dividing these individual SCFAs by total SCFAs. Total branched-chain fatty acids (BCFAs) were the sum of isobutyrate and isovalerate. The ratio of BCFAs to SCFAs, an indicator of shifts in protein fermentation relative to carbohydrate fermentation, was calculated by dividing total BCFAs by total SCFAs. Fecal pH and dry mass percentage were measured as previously described ^42^. Bile acid derivatives were measured in raw fecal material using ultrahigh performance liquid chromatography/multiple-reaction monitoring-mass spectrometry at the University of Victoria Genome BC Proteomics Centre (BC, Canada), as previously described ^82^.

#### Mechanistic markers of host-microbiome interactions

EDTA-treated plasma was analyzed by electrochemiluminescence immunoassay (ELICA; Meso Scale Discovery^®^, MD, USA) for inflammatory cytokines (TNF-α, IL-6, IL-8, and IL-10) using the V-PLEX^®^ Proinflammatory Panel 1 Kit (Catalog Number K151A9H; intra-assay coefficient of variation 4.5%, 4.7%, 6.6%, and 6.3%, respectively). A single-plex assay was used to measure adiponectin (Catalog Number K151BXC; CV 3.7%). Inhibitor-treated plasma was analyzed for metabolic hormones using the V-PLEX^®^ Metabolic Panel 1 Kit to measure glucagon, GLP-1, and leptin (Catalog Number K15174C; CV 4.5%, 5.0%, and 5.0%, respectively). The U-PLEX^®^ Metabolic Group 1 assay was also used to measure ghrelin and PYY (Catalog Number K151ACL; CV 4.2% and 4.0%, respectively). All assays were analyzed using the SECTOR^®^ Imager 6000 or MESO QuickPlex^®^ SQ 120 according to manufacturer protocols. Markers of gut barrier function were assessed in EDTA-treated plasma (LBP) and raw fecal material (zonulin and calprotectin), as previously described ^23^. TMAO was quantified in serum by high performance liquid chromatography-tandem mass spectrometry at the University of Alberta, as previously described ^82^.

#### Risk markers of chronic diseases

Height, measured to the nearest 0.01 cm (Digi-kit digital stadiometer, Measurement Concepts, Quick Medical^®^, WA, USA), and weight, measured using a calibrated digital scale to the nearest 0.1 kg (Health-o-Meter^®^ Professional 752KL, Pelstar LLC, IL, USA), were used to calculate BMI (kg/m^2^). Waist circumference was measured to the nearest 0.1 cm using a Gulick II plus tape measure (Country Technology Inc., WI, USA). Body fat percentage was estimated via bioelectrical impedance analysis using the Tanita^®^ Body Composition Analyzer (TBF-300A) and its proprietary equation (Tanita^®^ Corporation, IL, USA). Systolic pressure, diastolic pressure, and heart rate were measured using an automatic sphygmomanometer (Welch Allyn, Hill-Rom Inc., IN, USA). Inhibitor-treated plasma was analyzed to measure insulin by ELICA using the V-PLEX^®^ Metabolic Panel 1 Kit (Catalog Number K15174C; CV 4.3%). CRP was measured in EDTA-treated plasma using the V-PLEX^®^ Human CRP Kit (K151STD; CV 2.7%). All assays were analyzed using the SECTOR^®^ Imager 6000 or MESO QuickPlex^®^ SQ 120 according to manufacturer protocols.

Serum was used to quantify glucose and a lipid panel (total cholesterol, HDL cholesterol, non-HDL cholesterol, and triglycerides), obtained on a Beckman Coulter DxC 800 (Beckman Coulter Inc., CA, USA). Serum concentrations of LDL cholesterol were calculated using the Friedewald equation ^87^. Fasting glucose and insulin data were used to calculate HOMA-IR ^88^ and QUICKI ^89^.

#### Dietary intake

To assess if participants altered their dietary intake during the study, 24-hour recalls were administered using the Canadian version of the Automated Self-Administered 24-hour Dietary Assessment Tool (ASA24-Canada) (National Cancer Institute, MD, USA). Participants completed two consecutive recalls, one on a weekday and one on a weekend day, the data of which was averaged and used for statistical analyses. To characterize habitual diet in the previous month, participants also completed an online validated Canadian Diet History Questionnaire II (National Cancer Institute, MD, USA) ^90^. A Healthy Eating Index score adapted to Canada’s Food Guide (2007) was then calculated based on this questionnaire using the method outlined in Jessri et al. ^91^.

#### Gastrointestinal symptoms

A gastrointestinal symptom questionnaire ^43^ was used to determine the intensity of abdominal pain, bloating, flatulence, and overall symptoms using a 5-point hedonic scale, with no symptoms (0) and severe symptoms (4) on either end. Markers of gut transit time – self-reported stool consistency and bowel movement frequency – were also assessed in this questionnaire using a 5-point hedonic scale ^42^. The stool consistency scale had “hard or fragmented” (0) and “runny or watery” (4) on either end, with “smooth, soft, and formed” in the center (2). The bowel movement frequency scale had “every third day or less often” (0) and “three times a day or more often” (4) on either end, with “once a day” in the center (2).

Perceived satiety was assessed using the validated SLIM questionnaire ^47^ at three timepoints during each assessment day: 1) within 30-60 minutes of waking but before eating, 2) 30-60 minutes after eating a meal with the assigned fibre supplement added (at baseline, the meal had no fibre added), and 3) 2-2.5 hours after the same meal but before eating again. The SLIM questionnaire was a 100 mm, bidirectional hunger-fullness scale with “neither hungry nor full” in the center (0 mm) and had “greatest imaginable fullness” (50 mm) and “greatest imaginable hunger” (-50 mm) at either end. Participants drew a line across the scale to indicate perceived hunger or satiety intensity. The mark was measured from the center (0 mm) with a ruler and the distance was doubled to calculate the perceived satiety score. To assess differences in satiety scores between groups over the entirety of the intervention, the area under the curve was calculated using the linear trapezoidal method.

### Bioinformatics analysis

16S rRNA gene amplicon sequencing data were processed using QIIME2 ^92^, following our established pipeline ^23^ with one modified setting: to estimate alpha-diversity indices and the Bray-Curtis dissimilarity matrices among samples, the number of bacterial sequences was rarefied to 9000 reads per sample, and samples with read numbers<9000 were removed for further analyses. For the WMS dataset, the quality control steps were performed using our pipeline established previously ^23^. The compositional and functional profiles of the WMS dataset were generated using two complementary approaches: reference-mapping-based profiling (*i.e.*, reference-based) and *de novo* assembly-based profiling (*i.e.*, MAG-based). For reference-based analyses, post-QC unassembled short reads were analyzed using MetaPhlAn 3.0 with default parameters ^93^ to obtain relative abundances of bacterial and archaeal species. Microbial metabolic pathways were predicted based on short reads using HUMAnN 3.0 with default settings^93^. For MAG-based analyses, the same WMS dataset was used to generate *de novo* MAGs using the metagenome-atlas pipeline (version 2.17.1) ^94^, with default parameters. Briefly, post-QC reads were assembled into contigs using metaSPAdes (version 3.15.5) ^95^ and binned using VAMB (version 3.0.2) ^94^. High quality MAGs were selected based on the following criteria derived from CheckM2 (version 1.0.1) ^96^ quality assessment: (Completeness - 5 * Contamination > 50) & (Length of scaffolds ≥ 50000 nucleotides) & (Ambiguous bases < 1000000) & (N50 > 5000) & (N scaffolds < 1000). MAGs were then dereplicated at a 0.95 ANI (average nucleotide identity) threshold. CAZymes were annotated for the remaining MAGs using the dbcan2 implementation in DRAM 1.4.6, and CAZyme abundances were derived from MAG abundances, which were calculated as the median of the coverage of mapped reads in 1kb windows across each MAG. To quantify glycan-targeted CAZyme abundances, we grouped and summed the relative abundances of CAZymes according to their predicted or known substrate specificities ^97^.

To predict metabolic potential of microbial taxa identified by metagenomic analysis we used a subsystem-based approach implemented in microbial community SEED, an application of the SEED genomic platform ^98^. Metabolic subsystems curated across the representative human gut microbiome genomes include >100 biochemical pathways classified into three major categories: (i) biosynthesis of vitamins ^99^ and amino acids ^100^, (ii) production of SCFAs and other fermentation products^101^, (iii) utilization of carbohydrates, amino acids and other carbon sources ^102^. For each reconstructed metabolic pathway in each reference genome, we assigned a binary metabolic phenotype according to gene-specific pathway rules. The obtained Binary Phenotype Matrix (BPM) for reference genomes reflects the inferred presence or absence of respective 158 metabolic phenotypes. For functional annotation of MAGs and assignment of their binary metabolic phenotypes, we used this reference BPM and the computational pipeline Phenotype Predictor, as previously described ^103^. To calculate Community Phenotype Index (CPI), we used the obtained BPM for MAGs and their relative abundances in each metagenomic sample, as previously described ^104^.

Singleton and doubleton microbial features were excluded from the data derived from 16S rRNA gene amplicon sequencing prior to calculating α-diversity indices (*i.e.*, observed features, Shannon index, and Pielou’s evenness) and the β-diversity matrix (*i.e.*, Bray-Curtis dissimilarity); all other datasets did not have this filtering applied before calculating diversity indices. The α-diversity indices and β-diversity matrices were then estimated using the R package vegan ^105^. All features were filtered to retain only those present in at least 20% of the samples at a minimum of 0.1% of relative abundance for the downstream differential abundant feature identification.

### Statistical analyses

Analyses were performed using R v4.1.3 and GraphPad Prism v10.1.0. Only data from participants who completed the six-week intervention were included; for missing data, the participant(s) was removed for analyses of that variable. Due to the lengthy timeframe (almost five years) of the study and sample collection, samples had to be analyzed in two batches for bile acids, mechanistic markers of host-microbiome interactions, and some risk markers of chronic diseases (CRP, insulin, and calculations derived from insulin [HOMA-IR and QUICKI]). To account for potential batch-effects, a correction procedure was applied to these outcome variables ^106^. Briefly, for a given variable, the means of the two batches were calculated and divided by each other to determine the fold difference between the batches. Individual data points derived from the batch with the higher mean were then divided by the calculated fold difference to bring data on the same scale as the other batch. After this correction, the data no longer separated by ‘batch’. To validate this adjustment method, the unadjusted data was also analyzed by including ‘batch number’ (*i.e.,* processing group 1 or 2) included as a covariate in all statistical analyses ^107,108^, and results were virtually identical to analyses that used the batch-adjusted data (data not shown). Therefore, the batch-adjusted data was used for analyses and is presented in tables and figures. Data for the mechanistic markers and risk markers are presented as median (interquartile range [IQR]) as most were non-normally distributed.

For SCFAs, mechanistic markers of host-microbiome interactions, and risk markers of chronic diseases, outliers were removed based on a mean±5SD cut-off ^82^. For CRP, outliers were also identified for values >10 mg/L, likely indicative of infection ^109^. For 24-hour dietary recall data, outliers were identified using established cut-off points derived from NHANES data ^110^. Data distribution was assessed via visual inspection of histograms and Q-Q plots, and homogeneity of variance was examined using Levene’s test. Non-normally distributed data were transformed using either logarithmic or square root transformation. ‘Sex’ was included as a covariate due to participant stratification by this factor. To assess within-group changes, linear mixed models were applied using the *lme* function in the *nlme* package ^111^ to examine the effects of treatment (fibre assignment), time (baseline vs week 6), and their interaction, with participant ID included as a random effect. To assess between-group differences, analysis of covariance (ANCOVA) models were applied using the base *aov* function and Type III sum of squares. In addition to sex, the baseline value of the outcome variable was also included as a covariate, which was confirmed to be independent of treatment (fibre assignment) using the base *aov* function. When a significant main effect for ‘time’ or ‘treatment’ was detected in the linear mixed models and ANCOVA models, respectively, multiple comparison tests were conducted using the emmeans package ^112^ with Benjamini-Hochberg correction applied and FDR-adjusted *p*<0.05 considered significant (two-sided).

For perceived satiety, to determine within-group changes, scores derived from the SLIM questionnaire were assessed in the same way as above, with specific contrasts set to compare each weekly score to baseline. To assess between-group differences, calculated AUC values were used instead, with ANCOVA models applied as above. Since data derived from the gastrointestinal symptom questionnaire were ordinal and non-normally distributed, the *geepack* package was used for generalized estimating equations with repeated-measures models to assess the effect of fibre treatment over the six-week intervention ^113^.

Since bile acid data were also non-normally distributed, paired Wilcoxon tests were applied to assess within-group shifts compared to baseline, while unpaired Wilcoxon tests were applied for between-group comparisons at week 6. To correct for multiple comparisons, Benjamini-Hochberg correction was applied and FDR-adjusted *p*<0.05 was considered significant (two-sided). This approach was also applied to non-normally distributed microbiome data (taxonomic features, pathways, CAZymes, CPIs, and diversity indices), wherein paired Wilcoxon tests were used to assess within-group shifts and unpaired Wilcoxon tests were applied for between-group comparisons in AG or RS4 with MCC. A stricter cut-off was used for microbiome features, with FDR-adjusted *p*<0.01 considered significant (two-sided).

### Machine learning

The primary objective of the study was to determine if baseline microbiome features could predict physiological responses to the fermentable fibres (AG and RS4). To do so, separate random forest classifiers were trained based on taxonomic composition (ASVs from 16S rRNA gene amplicon sequencing and species from WMS), microbial pathways, CAZymes, aggregated CAZyme abundances, diversity indices, CPIs, SCFAs, and bile acids. The predictive value of baseline host features, namely dietary intake and immunometabolic status (*i.e.,* baseline values of all mechanistic and risk markers) was also evaluated. Random forest classifiers were chosen since they are particularly well-suited for high-dimensional, sparse, and potentially correlated features like those derived from microbiome data ^81,114^.

The machine learning procedure is depicted in the results in **Fig. S6.** Since many of the risk markers of chronic diseases were highly correlated with each other (**Fig. S6a**), representative markers of obesity (BMI), hypertension (diastolic blood pressure), dysglycemia (HOMA-IR), dyslipidemia (total cholesterol), and systemic inflammation (CRP) were chosen based on their highest correlation coefficient with other risk markers in these categories. Within the AG and RS4 groups separately, participants were divided into three equal tertiles based on their percent changes in these risk markers, and classified as either ‘responders’ or ‘non-responders’ depending on whether they were in the bottom tertile (*i.e.,* reduction in the marker) or top tertile (*i.e.,* increase in the marker), respectively (**Fig. S6b**). The participants in the middle tertile were excluded as their responses tended to hover around 0% change, and including them with the other ‘Non-Responders’ would result in unbalanced classes.

Independent classifiers were developed using 10-fold cross validation (**Fig. S6c**) using the default parameters in the *caret* package ^115^. To ensure an equal number of ‘Responders’ and ‘Non-Responders’ in each training and test set, the *fold* function was used from the *groupdata2* package ^116^. This procedure was repeated an additional four times for five-times repeated external cross validation due to the lack of a sizeable independent test set. To evaluate overall performance, the cross-validated area under the receiver operating characteristic curve (AUROC) was calculated using the *cvAUC* function in the *cvAUC* package ^117^. Classifiers with AUROC values >0.70 were considered to have good prediction accuracy. Mean feature importance for the top predictive features was calculated using the *varImp* function in the *caret* package.

Features from the individual datasets (MAGs, CAZymes, aggregated abundances of CAZymes, pathways, diversity metrics, CPIs, SCFAs, and bile acids, as well as metabolic status and dietary intake for baseline input) were also pooled to determine if combining microbiome- and host-derived features would improve predictive performance. All features were first correlated with each other using the base package *stats,* then hierarchical clustering was applied using the package *cluster* ^118^ with a threshold of *r* = 0.75. In each cluster, the feature with the highest mean correlation with its neighbours was kept (*i.e.,* representative feature), while all other features in that cluster were removed. The representative features from all clusters were then used as input for predictive models (named *‘All features combined’* in the Supplement).

To elucidate potential mechanistic underpinnings of the clinical effects, a similar machine learning approach was applied as above but used the absolute shifts in the input features instead of their baseline values. For these analyses, only those features that were significantly altered by either of the fermentable fibres were included as input. We further determined if responses in mechanistic markers of host-microbiome interactions could be predicted, as these outcomes are of additional interest in clinical applications. Only outcomes that were significantly altered by the fibres were selected for this analysis, and the same machine learning procedures were applied as above (using both baseline and shifts in predictive features), except that ‘Responders’ and ‘Non-Responders’ were defined based on the cohort median since there were few participants that had 0% change.

## DATA AND CODE AVAILABILITY

All original sequencing data (16S rRNA gene amplicon sequencing and whole metagenome sequencing) has been deposited at the NCBI Sequence Read Archive (SRA; accession number: SRA: PRJNA1301344). All other data can be made available on request pending written application and approval. No original or custom code was developed for this study. Researchers interested in the analytical code can contact the corresponding author.

## ACKNOWLEDGEMENTS

We would like to thank the participants for their involvement in this study. The intervention trial and participant clinical measures were completed at the Human Nutrition Research Unit, Department of Agricultural, Food and Nutritional Science at the University of Alberta. We thank Inés Martínez (Sacco Srl) for making conceptual contributions to product development and grant application. The analysis of whole metagenome sequencing (WMS) data was enabled by the support provided by Compute Ontario (computeontario.ca) and the Digital Research Alliance of Canada (alliancecan.ca). We thank Agrigum^®^ International Limited for providing the acacia gum fibre, MGP^®^ Ingredients for providing the resistant starch type 4 fibre, and Blanver Farmoquimica LTDA for providing the microcrystalline cellulose fibre.

This work was supported by the Canadian Institutes of Health Research (CIHR), Project Grant (PJT-152994). Bioinformatic analysis of the WMS dataset was in part supported by Nestlé Research. Funding for the measurements of fecal dry mass and gut barrier function biomarkers was provided by the German Federal Ministry for Education and Research (BMBF; grant no. 01EA1701) as part of the FiberTAG project (www.fibertag.eu), which was initiated by the European Joint Programming Initiative (JPI) “A Healthy Diet for a Healthy Life” (www.healthydietforhealthylife.eu). A.L.O., D.A.R. and A.A.A. acknowledge funding support from the National Institutes of Health (2R01DK030292-39). A.M.A. acknowledges funding support from the Izaak Walton Killam Memorial Scholarship, the Alberta Innovates Graduate Student Scholarship, the Frederick Banting and Charles Best Canada Graduate Scholarship, the Walter H. Johns Graduate Fellowship, and the University of Alberta Doctoral Recruitment Scholarship. F.L. acknowledges funding support from the “Hundred Talents Program” Research Start-up Fund of Zhejiang University, the National Natural Science Foundation of China, and the Alberta Innovates Postgraduate Fellowship. E.C.D. acknowledges support from the Anthony Fellowship in Human Nutrition, Elizabeth Russell MacEachran Scholarship, Queen Elizabeth II Graduate Scholarship, Frederick Banting and Charles Best Canada Graduate Scholarship, and Walter H. Johns Graduate Fellowship. C.M.P. acknowledges support through the Campus Alberta Innovates Program (CAIP) and Canada Research Chairs Program. J.W. acknowledges funding through CAIP, the Research Ireland Center grant to APC Microbiome Ireland (APC/SFI/12/RC/2273_P2), and a Research Ireland Professorship (19/RP/6853). The funding sources had no role in study design, data collection and analysis, decision to publish, or manuscript preparation.

## AUTHOR CONTRIBUTIONS

Conceptualization: E.C.D. and J.W. Funding acquisition and study design: E.C.D., J.A.B., J.M.C., C.J.F., S.D.P., W.V.W., C.M.P., D.K., J.W. Participant recruitment and data collection: E.C.D., A.M.A., and J.L.C. Clinical oversight of trial: A.M.S. Supervision: C.M.P. and J.W. Perceived satiety assessment: A.M.A., E.C.D., J.L.C., and W.V.W. Collection, DNA extraction, and chemical analysis of fecal samples: A.M.A., J.L., E.C.D., and F.L. Measurement of fecal dry mass and gut barrier function markers: B.S. and S.C.B. Measurement of cytokine and hormonal measures: A.M.A., E.C.D., and C.J.F. TMAO quantification: A.M.A., Y.Z., and J.M.C. Measurement of risk markers of chronic diseases: A.M.A., E.C.D., J.L.C., S.D.P., and C.J.F. Bioinformatics analyses of gut microbiome: D.N., L.S., and F.L. Carbohydrate-active enzyme analysis: O.D.B., L.S., B.B., and K.M.C. Assessment of community phenotype indices: D.A.R., A.A.A., and A.L.O. Statistical analyses: A.M.A., D.N., and J.A.B. Machine learning: A.M.A., R.G., and D.K. Writing of original draft: A.M.A., F.L., and J.W. Data interpretation and final manuscript: All authors.

